# Proteomic predictors of individualized nutrient-specific insulin secretion in health and disease

**DOI:** 10.1101/2023.05.24.23290298

**Authors:** Jelena Kolic, WenQing Grace Sun, Haoning Howard Cen, Jessica Ewald, Jason C. Rogalski, Shugo Sasaki, Han Sun, Varsha Rajesh, Yi Han Xia, Renata Moravcova, Søs Skovsø, Aliya F. Spigelman, Jocelyn E. Manning Fox, James Lyon, Leanne Beet, Jianguo Xia, Francis C. Lynn, Anna L. Gloyn, Leonard J. Foster, Patrick E. MacDonald, James D. Johnson

## Abstract

Population level variation and molecular mechanisms behind insulin secretion in response to carbohydrate, protein, and fat remain uncharacterized despite ramifications for personalized nutrition. Here, we define prototypical insulin secretion dynamics in response to the three macronutrients in islets from 140 cadaveric donors, including those diagnosed with type 2 diabetes. While islets from the majority of donors exhibited the expected relative response magnitudes, with glucose being highest, amino acid moderate, and fatty acid small, 9% of islets stimulated with amino acid and 8% of islets stimulated with fatty acids had larger responses compared with high glucose. We leveraged this insulin response heterogeneity and used transcriptomics and proteomics to identify molecular correlates of specific nutrient responsiveness, as well as those proteins and mRNAs altered in type 2 diabetes. We also examine nutrient-responsiveness in stem cell-derived islet clusters and observe that they have dysregulated fuel sensitivity, which is a hallmark of functionally immature cells. Our study now represents the first comparison of dynamic responses to nutrients and multi-omics analysis in human insulin secreting cells. Responses of different people’s islets to carbohydrate, protein, and fat lay the groundwork for personalized nutrition.

**ONE-SENTENCE SUMMARY:** Deep phenotyping and multi-omics reveal individualized nutrient-specific insulin secretion propensity.

## INTRODUCTION

Insulin is released by pancreatic islet beta cells in response to nutrient stimuli to maintain energy homeostasis. The major driver of insulin secretion is glucose. However, proteins and fats may also modulate insulin release and the effects of non-carbohydrate nutrients on insulin secretion remain underexplored. Much of our limited understanding of nutrient-stimulated insulin secretion is extrapolated from rodents, although more recent availability of cadaveric human islets for research purposes has expanded our pre-clinical knowledge. However, current human islet datasets generally only examine a single nutrient stimulus, glucose^1–3^. A few small studies examined other nutrients^4,5^, but no large-scale direct comparison of insulin secretion in human islets, stimulated by carbohydrates, proteins and fats has been reported. Understanding of nutrient-induced insulin secretion is important in the context type 2 diabetes and emerging studies linking hyperinsulinemia with cancer^6^. Indeed, large prospective clinical trials shows broad beneficial effects of diets targeting hyperinsulinemia^7^.

Individuals respond differently to diets^8^ and there is high interpersonal variability in postprandial responses to even one macronutrient, glucose^9^. *Ex vivo studies* measuring insulin secretion from human islets show high variability, only some of which can be explained by donor characteristics or islet isolation parameters^2^. The concept that insulin-responses to food types or different macronutrients are individualized has not been investigated. No studies have leveraged macronutrient-induced insulin secretion heterogeneity and large-scale multi-omics to elucidate the associated molecular mechanisms.

Here we address these critical knowledge gaps by measuring dynamic insulin secretion in response to three model macronutrient stimuli in islets from non-diabetic and donors with type 2 diabetes, as well as stem cell-derived islets. Our transcriptomic and proteomic analysis revealed molecular signatures of each islet donor and identified distinct clusters of proteins that predict insulin secretion responses to carbohydrates, proteins, and fats. We also report on a subset of donor islets that islets that are relatively hyperresponsive lipids in a way that resembles functionally immature human embryonic stem cell-derived islets. This is the largest human islet dataset that includes both macronutrient-stimulated insulin secretion measurements and multi-omic profiling, coupled with the first side-by-side comparison of nutrient responses and proteomes between human islets and stem cell-derived islet clusters. This resource helps us understand why how individuals’ islets respond differently to sugar, protein, and fat and advocates for greater application of personalized recommendations and treatments for individuals living with diabetes.

## RESULTS

### Prototypical nutrient-stimulated insulin secretion dynamics from human islets

Between 2016 and 2022, we systematically measured insulin secretion in response to carbohydrate (15 mM or 6 mM glucose), amino acid (5 mM leucine), and fat (1.5 mM oleate/palmitate mix) from islets isolated from 140 cadaveric donors^2^ reflective of the general population (Fig. 1A)^10^. In most islet donors, we confirmed that carbohydrate was the strongest insulin secretagogue, followed by amino acid and then fat, which only weakly stimulated insulin secretion on average (Figs. 1B, S1A,B). Islets exhibited biphasic insulin secretion in response to glucose^11^. We found that insulin secretion in response to amino acid is also biphasic, with a distinct 1^st^ phase lasting ∼15 minutes and a sustained 2^nd^ phase lasting the duration of the challenge (Figs. 1B, S1A,B). In contrast, the response to fat, when present, was monophasic (Figs. 1B, S1A,B).

**Fig. 1.**
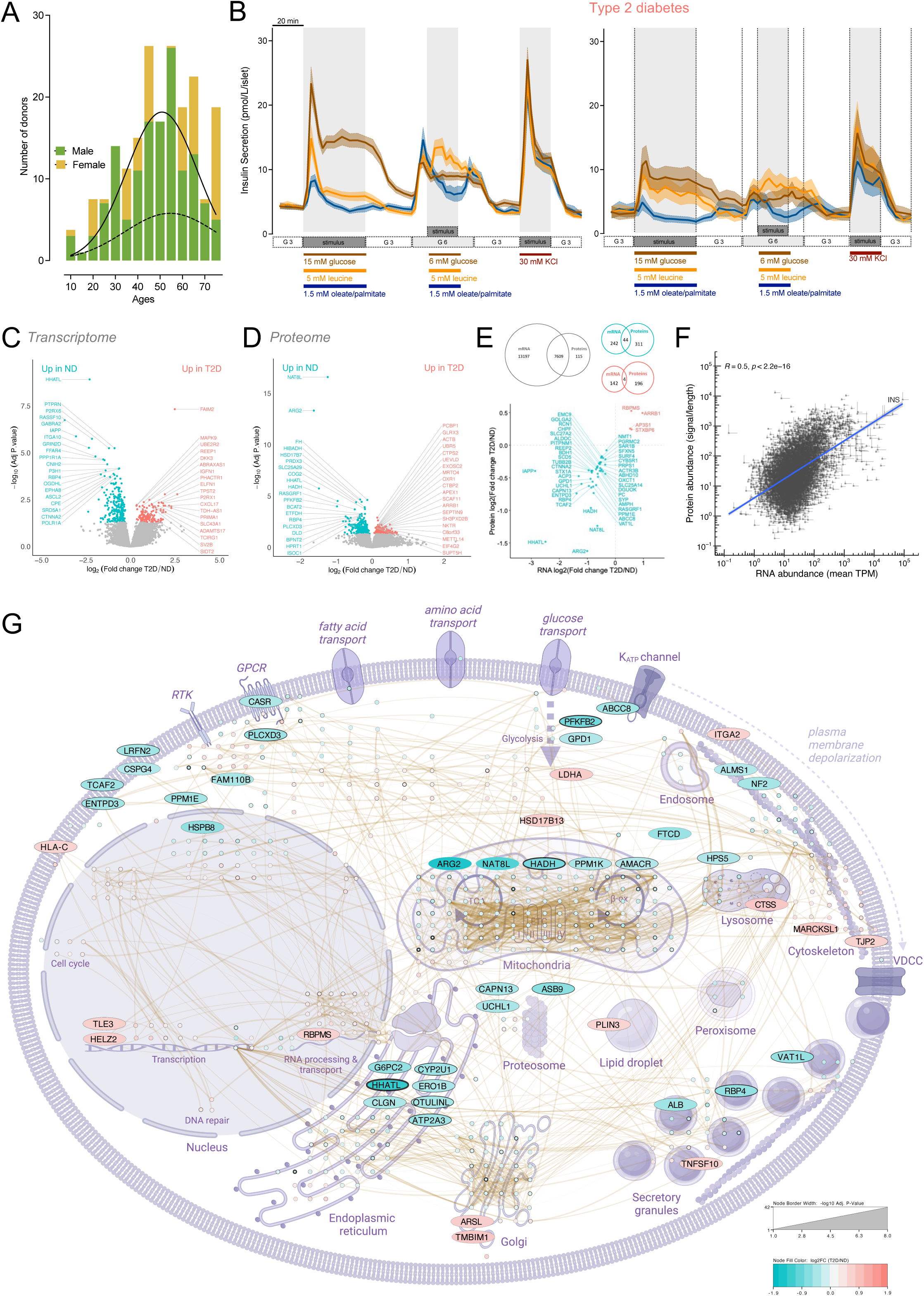
Nutrient-stimulated insulin secretion, transcriptomes, and proteomes from metabolically healthy and T2D islets. (**A**) Histogram with trend line of ages for both male (green, solid line) and female (yellow, dashed line) donors. Additional details of islet isolation parameters and donor characteristics are available in Table S1 and will be available on www.humanislets.com **(B)** Averaged traces of dynamic insulin secretion measurements in response to glucose (15 or 6 mM), leucine (5 mM), 1.5 mM oleate and palmitate (1:1 mixture) or 30 mM KCl in islets isolated from non-diabetic donors (n=123) (*left)* and donors with type 2 diabetes (n=17) *(right)* are shown as indicated. Basal glucose was 3 mM. **(C)** Islet samples from the cadaveric donors were collected for RNAseq. Enriched mRNAs in islets from ND donors (n= 82) are shown in teal and enriched mRNAs in islets from donors with type 2 diabetes (n=8) are shown in red. The top 40 most significant differentially abundant proteins are highlighted by labelling with gene name. **(D)** Shows the same as E, except donor islets were collected for mass-spec based proteomics (n= 118) in ND, and (n=16) in T2D **(E)** *(top)* Venn diagrams show the overlap of differentially expressed mRNAs and abundant proteins, and this is further depicted in the correlation plot *(bottom)* **(F)** Across-gene correlation between RNAs and proteins **(G)** Differentially abundant proteins (with a greater than 0.5 log2 fold change) between the non-diabetic and type 2 diabetic donor islets were connected using STRING and depicted in the context of beta cell signalling. The color of the nodes represents the fold change (ND/T2D), while the thickness of the line around the nodes represents the p-value (adjusted).

We also assessed the role of macronutrient order on insulin secretion, inspired by clinical meal-order studies^12,13^. In our experiments, prior exposure to high glucose, amino acid or fat did not alter insulin secretion stimulated by moderately elevated 6 mM glucose (Figs. S1A,B). Amino acid on top of 6 mM glucose further increased insulin secretion (Figs. S1A). Fat produced no enhanced response (Figs. S1A,B), in contrast to previous small-scale rodent or human islet observations^14,15^. Interestingly, insulin secretion stimulated by direct depolarization with KCl was inhibited after prior exposure to lipid in islets from normoglycemic donors (Fig. S1A), perhaps foreshadowing lipotoxic effects on the insulin secretory machinery^16^. This large dataset provides the first side-by-side response profiles for each of the main three macronutrients in human islets.

### Prototypical nutrient-stimulated insulin secretion dynamics in type 2 diabetes

We next examined the relationship between type 2 diabetes and nutrient-stimulated insulin secretion. Islets from donors with family-reported type 2 diabetes (Table S1) had ∼40% lower insulin secretion in response to 15 mM glucose and ∼35% lower insulin secretion in response to moderate 6 mM glucose (Figs. 1B, S2C,F). Insulin secretion in response to direct depolarization by KCl was reduced by ∼22% but failed to reach statistical significance (Fig. S2I, p=0.07). When examining the kinetics of insulin secretion, we saw a significant delay in time-to-insulin peak in response to high glucose in islets from donors with diabetes (Fig. S1E). Insulin secretion in response to fatty acids was also lowered by ∼55% (Figs. 1B, S2E). However, insulin content, baseline insulin secretion, and insulin secretion in response to 5 mM leucine were not different (Figs. 1B, S2A,B,D). This preserved amino acid-stimulated insulin secretion is consistent with clinical data^17^ and suggests therapeutic protein intake could be exploited in diabetes management. However, leucine together with 6 mM glucose induced ∼36% (on average) less insulin secretion in donors with diabetes (Fig. S2G), and leucine was not stimulatory on top of 6 mM glucose in these donors (Fig. S1B), emphasizing a need for additional clinical research into context-dependent amino acid-stimulated insulin secretion. Known glucose-lowering medication status of these donors with diabetes did not significantly correlate with any insulin-secretion parameters; however, there was an overall trend with the need for exogenous insulin and lower overall insulin secretory capacity (Fig. S2J-Q).

### Comprehensive transcriptomics and proteomics of islets with and without type 2 diabetes

To better understand the relationship between secretory response and variation in gene expression in islets from donors with and without type 2 diabetes, we performed comprehensive transcriptomic and proteomic analysis from corresponding batches of donor islets. On average, after quality control, we measured over 20,000 mRNAs using RNA sequencing (from 82 ND and 8 T2D donors) and almost 8000 proteins (from 118 ND and 16 T2D donors) using mass-spectrometry. We first used our multi-omic dataset to estimate islet cell composition in our donor preps based on abundance of proteins typically enriched in islet and acinar cells^18^. The model indicates that islets from donors with T2D have altered islet cell composition, with lower beta cell and higher alpha and delta cell markers, as well as a higher percentage of acinar cell markers (Fig. S3B,D).

Using RNA sequencing of a large subset of human islets, we next showed that 286 mRNAs were enriched in the non-diabetic donors and 146 were enriched in the donors with diabetes (Fig. 1C, Table S2). Importantly, differences were more apparent at the protein level, with 355 proteins significantly more abundant in islets from non-diabetic donors, and 200 proteins more abundant in islets from donors with diabetes (Fig. 1D, Table S3). Despite only finding 48 gene products that were significantly altered at both mRNA and protein levels (Fig. 1 E), individual protein fold changes were mostly consistent with mRNA fold changes (Fig. S4A), and both lists of features were enriched with many of the same pathways (Fig. S4B). We also used an alternative approach to compare the protein and RNA changes in T2D by utilizing the rank-rank hypergeometric overlap test^19^, which identified ∼3300 proteins and RNAs that were changed in the same direction in T2D (Fig. S5). However, we found that this method was not stringent enough for our purposes since the vast majority of the overlapping proteins and RNAs it identified had negligible fold change difference in T2D (Fig. S5).

Gene products decreased in both expression and abundance in islets from donors with diabetes included the sulfonylurea receptor subunit of the K_ATP_ channel (*ABCC8*), the well-known target of oral hypoglycaemic agents, as well as pyruvate carboxylase (*PC*), which is critical for mitochondrial metabolism and glucose-stimulated insulin release^20^, hedgehog acyltransferase-like (*HHATL*), which negatively regulates protein palmitoylation, a process implicated in type 2 diabetes^21^ and islet amyloid polypeptide (*IAPP*), a hormone co-secreted with insulin with roles in glycemic control and gastric emptying^22^ (Fig. 1E). Only four gene products were increased in both expression and abundance in type 2 diabetes: RNA-binding protein with multiple splicing (RBPMS), AP-3 complex subunit sigma-1 (AP3S1) – involved in lysosomal trafficking^23^, syntaxin binding protein 6 (*STXBP6*) which is thought to limit insulin release by limiting the size of the granule fusion pore^24^ and beta arrestin 1(*ARRB1*) which is suggested to be involved in GLP-1 stimulated insulin secretion^25^.

There was overall discordance between islet RNA expression and protein abundance (7609 gene products) from the same donor/isolations, with an R value of 0.5 (Fig. 1F), consistent with other studies of primary tissues^26,27^. As such, we did not detect significant RNA-protein correlations for 81% of the gene products, which interestingly mostly encoded for mitochondria-related and ribosomal proteins (Fig. S4C,D,E). This discrepancy was likely not due to unreliable RNAseq measurements because we observed a strong correlation between the RNAseq and NanoString analysis of 130 mRNAs (Fig. S4F,G). Because proteins had lower co-efficients of variation (Fig. S4H), were more stable in the face of isolation variables (see below), and may provide better insight into disease phenotype than mRNAs^27^, we focused on the proteomic data to elucidate key mechanisms. We mapped the protein-protein interaction networks for proteins showing fold-change greater than 0.5 log2 using STRING^28^, and subsequently assigned them to their intracellular location in a diagram using subcellular location information found in UniProt (Fig. 1G). Consistent with a lower insulin secretory response to stimulatory glucose (Fig. S2C,F), islets from donors with type 2 diabetes had lower abundance of key proteins predicted to be involved in glucose-stimulated insulin secretion. Islet proteins reduced in type 2 diabetes (Fig. 1 G) included mitochondrial proteins arginase 2 (ARG2), N-acetylaspartate synthetase (NAT8L), hydroxyacyl-coenzyme A dehydrogenase, mitochondrial (HADH), protein phosphatase 1K, mitochondrial (PPM1K) and alpha-methylacyl-CoA racemase (AMACR), regulators of glycolysis (6-phosphofructo-2-kinase/fructose-2, 6-bisphosphatase 2 (PFKFB2) and glucose-6-phosphatase 2 (G6PC2), endoplasmic reticulum Ca^2+^ homeostasis factors ERO1-like protein beta (ERO1B), inactive ubiquitin thioesterase (OTULINL), sarcoplasmic/endoplasmic reticulum calcium ATPase 3 (ATP2A3), and the extracellular Ca^2+^-sensing receptor (CASR). Proteins that were more abundant in islets from donors with diabetes included those crucial for cell adhesion (tight junction protein ZO-2 (TJP2), integrin alpha-2/beta-1 (ITGA2), Golgi function arylsulfatase L (ARSL), transmembrane BAX inhibitor motif containing 1 (TMBIM1), transcriptional proteins such as TLE family member 3 (TLE3), helicase with zinc finger domain 2 (HELZ2) and RNA-binding protein with multiple splicing (RBPMS), and proteins involved in cytoskeletal reorganization like MARCKS-related protein (MARCKSK1). Collectively, these data identify multiple proteins that correlate with the insufficient glucose-stimulated insulin secretion in islets from donors with diabetes.

### Individuality of nutrient-stimulated insulin secretion

Islets from individual donors exhibited a large range of insulin secretion rates at baseline, and in response to high glucose, moderate glucose, amino acid, fat, and direct depolarization (Fig. 2A). Surprisingly, we observed that some donors had more robust responses to fatty acids than to glucose, challenging the long-standing idea that dietary fats alone have negligible effects on insulin release^5,14,29^. This degree of response heterogeneity across all macronutrients was not found in C57Bl6J mouse islets, even when including both sexes and a wide range of ages (Fig. 2B).

**Fig. 2.**
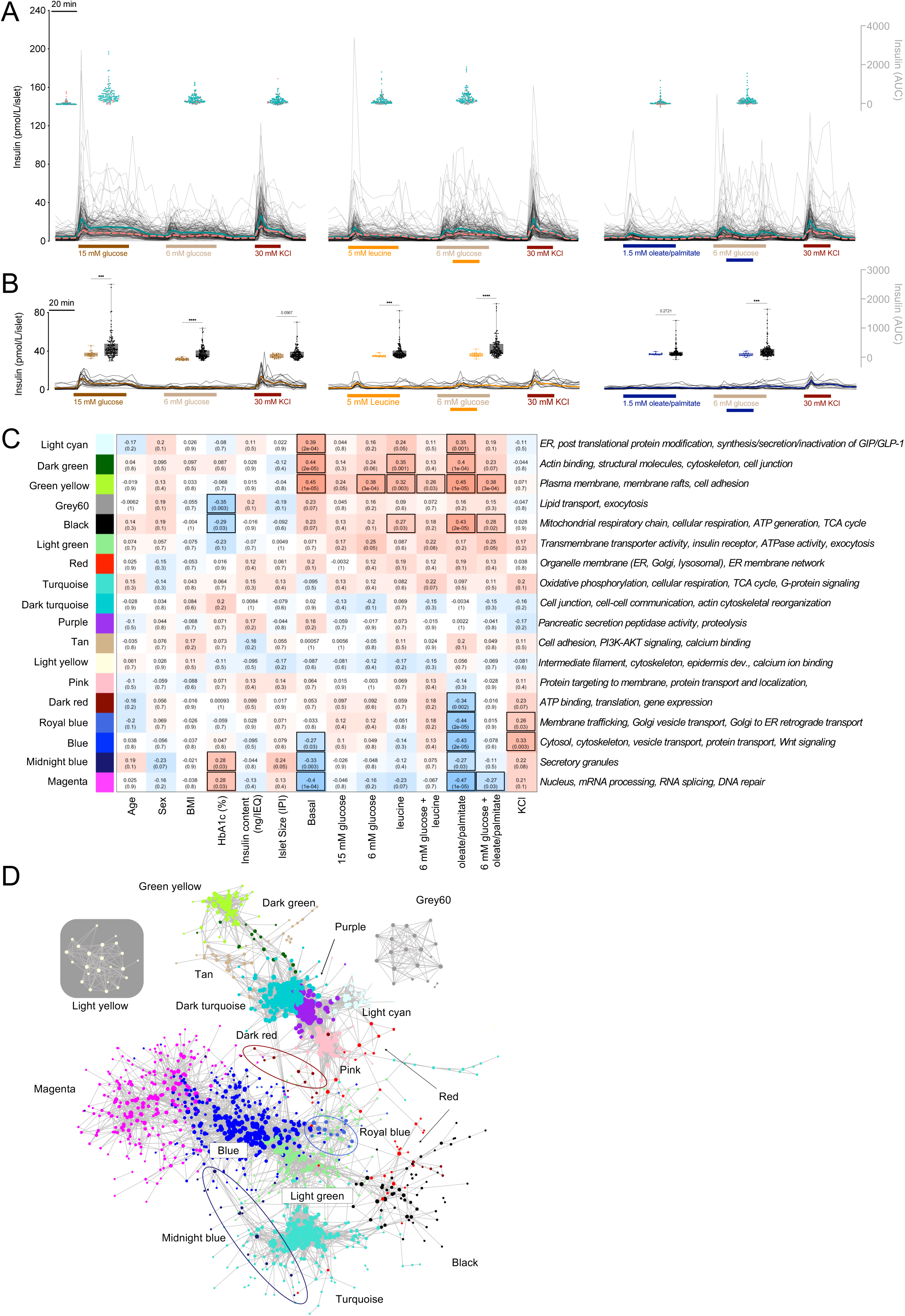
Clustering heterogenous insulin responses to macronutrients and donor proteomes. (**A**) *(left)* Individual traces of dynamic insulin secretion stimulated by glucose (15 mM or 6 mm) or KCl (30 mM). Basal glucose was 3 mM. Average responses from non-diabetic donors are illustrated with solid teal line and average responses from donors with type 2 diabetes are in shown in the dashed salmon line. Floating dot plot inserts illustrate the heterogeneity in insulin AUC for the corresponding section of the perifusion curve (salmon dots illustrate the responses from donors with T2D). *(middle)* illustrates the same as *(left)* except islets were stimulated with 5 mM leucine alone, or in combination with 6 mM glucose as depicted in figure panel. *(right)* illustrates the same as *(left)*, except islets were stimulated with 1.5 mM oleate/palmitate (1:1 mix), or in combination with 6 mM glucose. **(B)** Illustrates the same as (A) except in mouse islets of both sexes (7 males, 9 females, 7-90 weeks of age). Floating box plots illustrate the variance of AUC responses between the mouse islet (grey) and human islet (brown) responses. **(C)** Co-correlation analysis identified 18 distinct modules of proteins whose abundance shows similar patterns. Modules are annotated with KEGG pathways and GO terms. The Pearson correlation coefficients between the modules and the donor meta data or functional data (x-axis) are shown in each rectangle, and the corresponding adjusted p value is shown in the brackets. Positive correlations are shown in shades of red and negative in shades of blue. Significant correlations are highlighted with black boxes. **(D)** Illustrates the main connection in the co-expression network. The WGCNA adjacency matrix was filtered to remove: 1) any protein not in a module, 2) protein-protein adjacency distances less than 0.2, and 3) any protein without any connections after filters 1 and 2. The resulting network contained 1600 protein nodes and 16 209 edges. Each protein node is colored according to its module (same color scheme as C).

We explored the source of this variation by examining known donor characteristics (Table S1) and islet isolation parameters. Cold ischemic time of the pancreas was negatively correlated with insulin secretion in response to direct depolarization (Fig. S6P), but it was not correlated to insulin secretion stimulated by any of the three macronutrients tested. We also found that islets from female donors had lower insulin secretion at 3 mM glucose and 6 mM glucose, with or without fatty acid treatment (Fig. S6A-C). Donor age and BMI were not different between male and female donors in our study (Table S1), suggesting that these differences are a result of biological sex. Somewhat surprisingly, donor BMI had only minor effects on macronutrient-stimulated insulin secretion (Fig. S6D-L). Expectedly, high HbA1c was negatively correlated to insulin secretion stimulated by high glucose and KCl (Fig. S6M-O). These observations suggest that donor characteristics and islet isolation parameters minimally contribute to individualized nutrient responses and that separate factors must drive the observed heterogeneity.

### Co-expression network analysis uncovers protein-networks correlated with islet function

We used co-expression network analysis to obtain an overview of protein-protein relationships within the proteome (and transcriptome), and then analyzed the network along with clinical and functional outcomes to gain insight into the molecular drivers of macronutrient-stimulated insulin secretion response heterogeneity. We identified 18 network modules of highly co-regulated proteins (Figs. 2C, S7) and plotted them as a co-expression network to illustrate the connections between modules (Fig. 2D). All modules were significantly enriched in gene sets (Gene Ontology Cellular Component, Molecular Function, and Biological Process) and pathways (KEGG and Reactome), allowing for their higher-level functional annotation. The overall protein abundances of 9 modules were significantly correlated with islet functional data (Fig. 2C). Several modules were positively correlated with insulin secretion in response to multiple nutrients. These modules (lightcyan, dark green, greenyellow and black) contained proteins with critical roles in cytoskeletal reorganization, mitochondrial metabolism, and insulin processing. Two modules of interest (blue and royalblue) with roles in protein transport and localization, were positively correlated with insulin secretion stimulated by KCl but negatively correlated with insulin secretion stimulated by lipids. Notable proteins in this module include enzymes involved in fatty acid synthesis, acetyl-CoA carboxylase 1 (ACACA) and fatty acid synthase (FASN)^30,31^, and lipid metabolism glycerol-3-phosphate dehydrogenase (GPD1)^32^. Consistent with the discordance between RNA expression and protein abundance, and suggestive of more dynamic mRNA levels, we did not observe any significantly correlated transcript modules in the 29 network modules detected (Fig. S8). Together, these data demonstrate that these protein network modules in human islets likely underpin their differential responsiveness to stimuli.

### Associations between individual proteins, donor data and islet function

We first correlated the abundance of each mRNA (Fig. S9, Table S4) and protein (Fig. S10, Table S5) identified in our dataset to donor traits, independent of diabetes status. Some of these correlations were driven by disease status rather than islet function (Table S6), so we next reanalyzed our dataset using a linear regression model to adjust for T2D diagnosis (Fig. 3, S11). We focused our attention on the proteome because we found that mRNAs were poorly correlated with islet function (Fig. S9,11) and were more prominently associated with islet culture and isolation variables (Fig. S12, Tables S6-8).

**Fig. 3.**
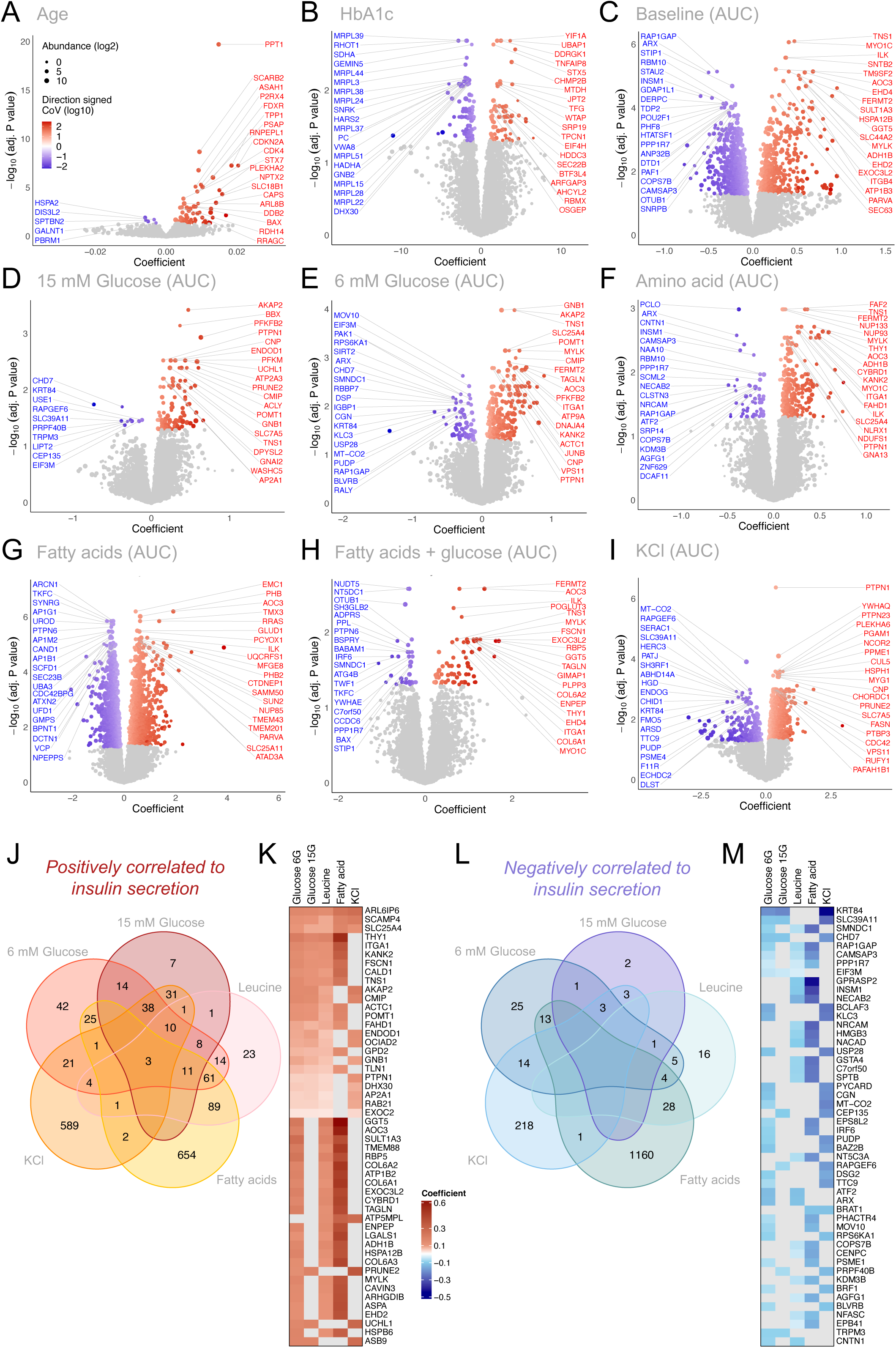
Regression coefficients (adjusting for T2D status) of individual proteins with donor characteristics and insulin secretory responses. Volcano plots are shown depicting significant positive (red) and negative (blue) linear regression coefficients of proteins to continuous donor characteristics: **(A)** donor age and **(B)** HbA1c; or functional parameters in response to: **(C)** 3 mM glucose **(D)** 15 mM glucose **(E)** 6 mM glucose **(F)** 5 mM leucine **(G)** 1.5 mM oleate/palmitate (1:1 molar ratio) **(H)** 1.5 mM oleate/palmitate + 6 mM glucose and **(I)** 30 mM KCl. Protein abundances (log2) are depicted by size of circle, and the coefficient of variation (log10) is depicted by color gradient. **(J)** Venn diagram showing the overlap of the number of protein abundances that positively associate with the indicated nutrient stimuli following adjustment for T2D disease status diagnosis **(K)** Heat map depicts the top 50 most positively associated proteins to the indicated stimuli. **(L-M)** Shows the same as (J-K), but for negative associations.

Our analysis suggests that increasing donor age was positively associated to proteins with known (and postulated) roles in lysosomal degradation, cellular senescence, inflammation and mitochondrial dysfunction: palmitoyl-protein thioesterase 1 (PPT1)^33^ scavenger receptor class B member 2 (SCARB2)^34^, acid ceramidase (ASAH1)^35^, purinergic receptor P2X4^36^, and NADPH:adrenodoxin oxidoreductase (FDXR)^37^ (Fig. 3A, Table S8). We found that the abundances of 161 proteins were associated with HbA1c (Fig. 3B). Interestingly, the most significant positive relationships were seen to proteins with known roles in cancer progression and development: protein YIF1A^38^, ubiquitin associated protein 1 (UBAP1)^39^, DDRGK domain-containing protein 1^40^ and syntaxin-5 (STX5)^41^. This is relevant to reports of poor glycemic control being associated with and increased risk of multiple cancers^42^.

We also looked for relationships between protein abundance and islet function in response to each nutrient stimulus (Fig. 3C-I). The greatest number of significant associations to protein abundance were seen with insulin secretion induced by lipids (2053 significant associations) (Fig. 3G, Tables S6,8), pointing to somewhat unique mechanisms for this macronutrient as a possible modulator of insulin release. The abundance of 3 proteins was positively associated to insulin secretion stimulated by all three macronutrients as well as direct depolarization by KCl (Fig. 3J,K), suggesting that these proteins play a role in the overall secretory capacity of the beta cell. These pan-stimulus enabling proteins were ADP-Ribosylation Factor GTPase 6 Interacting Protein 6 (ARL6IP6), solute carrier family 25 member 4 (SLC25A4), and secretory carrier membrane protein 4 (SCAMP4). These findings highlight the critical role of mitochondria in nutrient– and depolarization-stimulated insulin secretion^43^.

Interestingly, alpha cell transcription factor, aristaless related homeobox (ARX)^44^ was negatively associated with basal insulin secretion (Fig. 3C), insulin secretion stimulated by glucose (Fig. 3E), as well as insulin secretion stimulated by amino acid (Fig. 3F) (Table S8). While these correlations may initially suggest a general decrease in beta cell ability to secrete insulin on account of higher alpha cell mass, we also observe that insulin secretion induced by direct depolarization with KCl was not correlated to ARX abundance (Fig. 3M, Table S8), arguing against such a simple interpretation. Collectively, these data provide unprecedented information on the proteins that associate with islet secretory function, in a general and nutrient-specific way. We have built an online resource where the relationship between any detected protein, donor characteristics, islet isolation parameters and islet function can be displayed (www.humanislets.com).

### Unique proteome of lipid– or amino acid-hyper-responders

During the course of our multi-nutrient phenotyping studies, we identified a previously un-reported sub-group of donors with unusually large insulin secretory response to lipids in basal glucose conditions. In fact, ∼8% of donors secreted more insulin in response to oleate/palmitate than to 15 mM glucose (Fig. 4A). These donors also secreted more insulin when lipid was present together with 6 mM glucose, but less insulin in response to direct depolarization by KCl (Fig. 4A, Table S9). Overall, lipid hyper-responsive islets came from donors with higher HbA1C, but no significant differences in known islet isolation or culture parameters were seen between the two groups (Table S9).

**Fig. 4.**
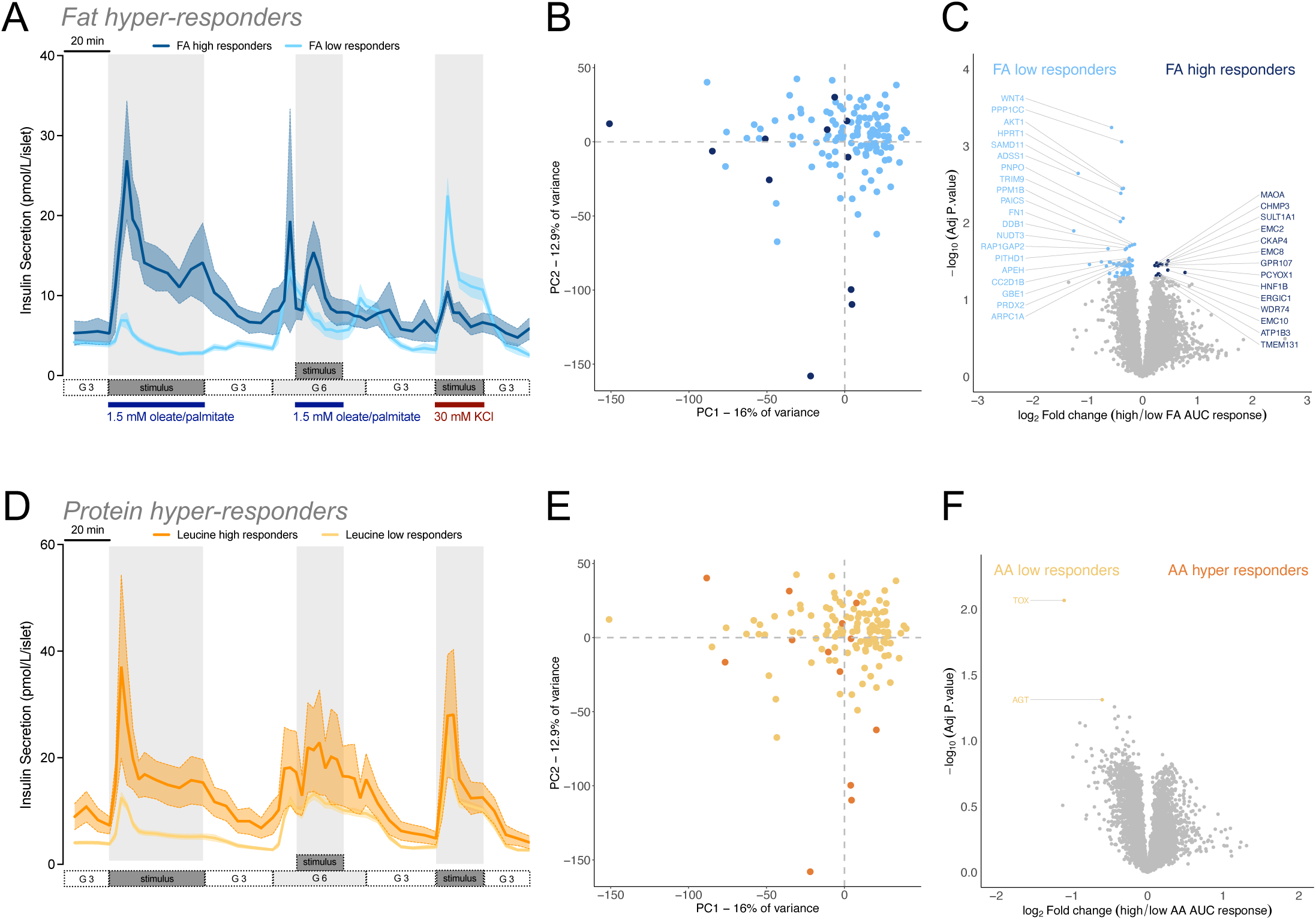
Prototypical proteomes and responses of fat and protein hyper-responders. (**A**) The average insulin secretion response from donors classified as “high lipid responders” is shown in dark blue (n=11) and the average insulin secretion response from donors classified as “low lipid responders” is shown in the light blue (n=129). **(B)** Principal component analysis (PCA) plot of protein abundance in the high (dark blue circles) vs. low (light blue circles) lipid responders. **(C)** Volcano plot showing differential protein abundance between high and low lipid responders. **(F)** The average insulin secretion response from donors classified as “high protein responders” is shown in orange (n=13) and the average insulin secretion response from donors classified as “low protein responders” is shown in yellow (n=127). **(G)** PCA plot of protein abundance in the high (orange circles) vs. low (yellow circles) protein responders. **(H)** Volcano plot showing differential protein abundance between high and low protein responders.

However, proteomic comparison of these two donor groups showed that the fat hyper-responsive islets exhibited a 30% decrease in the abundance of Wnt family member 4 (WNT4), hinting at a less-mature state^45^ (Fig. 4B,C). Fat hyper-responsive islets also had less protein phosphatase 1 catalytic subunit gamma (PPP1CC), which is involved in glycogen metabolism, hypoxanthine phosphoribosyltransferase 1 (HPRT1), which is involved in purine metabolism, and AKT serine/threonine kinase 1 (AKT1), which is involved in growth factor signaling and survival. On the other hand, lipid hyper-responders exhibited elevated hepatocyte nuclear factor 1-beta (HNF1B), the gene product underlying a rare form of diabetes, maturity onset diabetes of the young (MODY5), sulfotransferase family 1A member 1 (SULT1A1), and platelet derived growth factor receptor alpha (PDGFRA) (Fig. 4B,C). Pathway analysis using multiple databases ^46^ reveals defects in small-molecule protein modification, AKT signaling and lipid metabolism in donors classified as fat-hyper responders (Fig. S13A). Interestingly, proteins involved in endoplasmic reticulum signal integration and organization are increased in fat-hyper responders (Fig. S13B), which is perhaps suggestive of endoplasmic reticulum dysfunction in these donors^47^.

We also identified a previously un-reported subset of donors (∼9%) that secreted more insulin in response to leucine than to high glucose (Fig. 4D, Table S11). Amino acid hyper-responders were more likely to been diagnosed with type 2 diabetes, but there was no difference in HbA1C between the high and low responders (Table S10). Amino acid hyper-response was also associated with a longer *in vitro* culture time (Table S10), suggesting that this observed phenotype may be influenced by adaptation to our culture conditions. Only two differentially abundant proteins (Fig. 4E,F) were associated with this phenotype, thymocyte selection-associated high mobility group box protein (TOX; transcriptional regulator) and angiotensinogen (AGT), an essential component in the renin-angiotensin system, with important roles in the endocrine pancreas^48^. Our work now suggests that some donors’ islet proteomes may be pre-programmed or may have adapted to *in vitro* culture conditions, which enables them to hyper-respond to lipids and/or amino acids.

### Human embryonic stem cell-derived islet-like clusters are lipid-responsive

The differentiation of beta cell surrogates from embryonic stem cells has potential as a diabetes therapy and can be used to model ‘environmentally naïve’ insulin secretion^49^. The *in vitro* insulin-secretory function of these cells, even in response to supraphysiological levels of glucose, still does not match that of human islets^49^. There are no reports that directly compare how stem cell-derived islets and primary human islets respond to multiple nutrients^49^, however recent work has suggested that human mitochondria from pluripotent stem cell-derived islets have a higher sensitivity for non-glucose fuels^50^. In our hands, INS2AGFP stem cell-derived islet-clusters^51^ (Fig. 5A,B) exhibited lower levels of typical beta cell enriched markers (Fig. S3C,D) and had ∼1/10 the level of basal insulin release when compared to human islets (Fig. 5C). They showed poor responsiveness to glucose, were unresponsive to amino acid (either alone or in combination with glucose) but did release significant amounts of insulin upon direct depolarization by KCl (Figs. 5C, S14). This suggests that these *in vitro* derived cells have the capacity for regulated insulin secretion but have specific defects in glucose and amino acid responses and may prefer non-glucose substrates^50^.

**Fig. 5.**
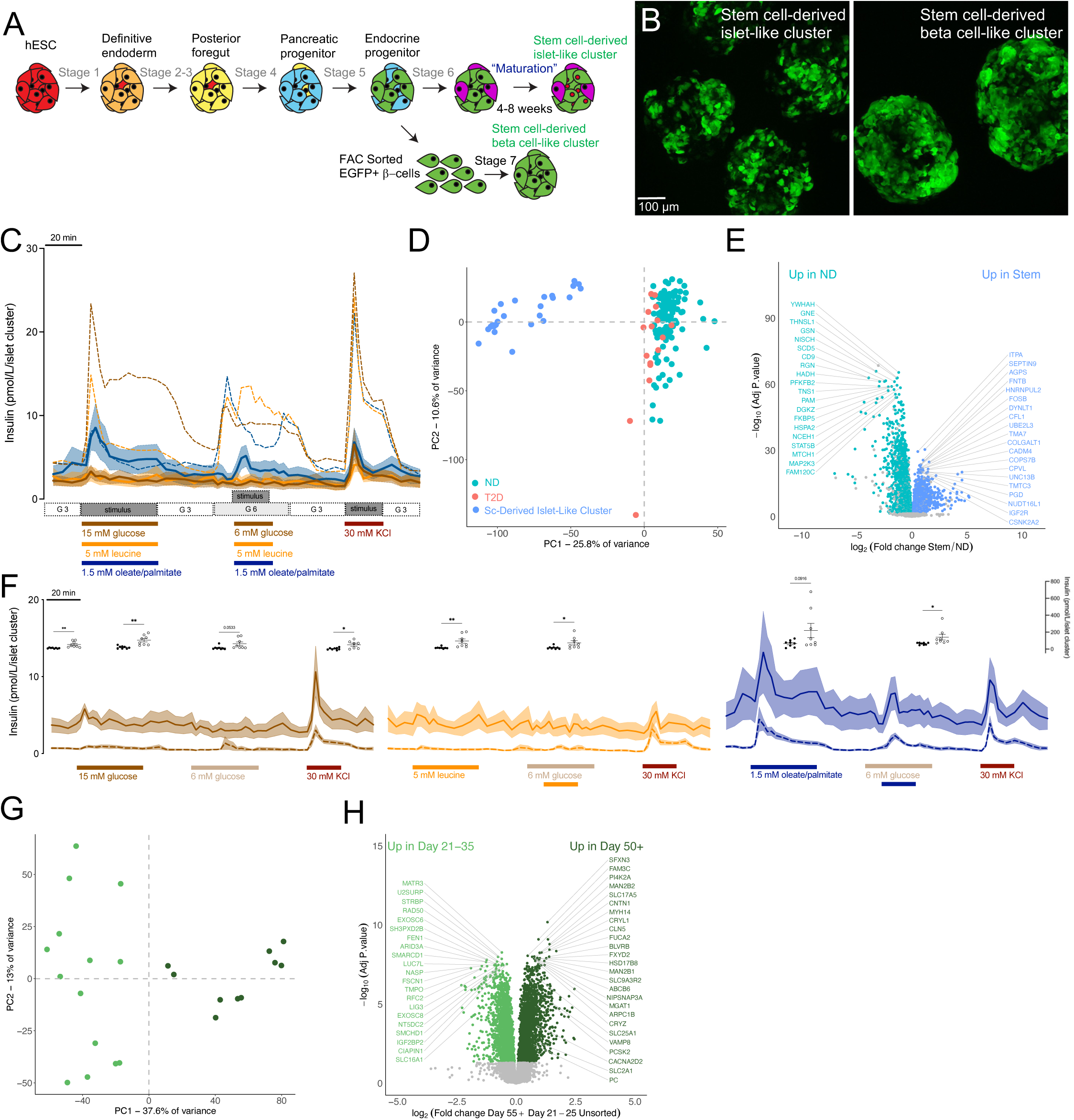
Stem cell-derived islet-like clusters hyper-respond to fat and can be matured with extended culture. (**A**) Summary of the human embryonic stem cell-derived islet-like clusters differentiation protocol. **(B)** Representative images of unsorted (day 35) Sc-derived islet-like clusters *(left)* and FAC-sorted (day 35) Sc-derived beta cell-like clusters *(right)* **(C)** Averaged traces of dynamic insulin secretion measurements in response to glucose (15 or 6 mM), leucine (5 mM), 1.5 mM oleate and palmitate (1:1 mixture) or 30 mM KCl in stem cell derived islet-like clusters (average of 8 younger-immature and 8 older-maturing preparations). Basal glucose was 3 mM. For comparison, the average human islet dynamic insulin secretion measurement traces are shown in the dashed lines. **(D)** PCA plot of protein abundance in the stem cell-derived islet-like clusters (n=25) (blue circles) compared to donors with type 2 diabetes (salmon circles, n=16) and normoglycemic donors (teal circles) (n=118). **(E)** Volcano plot showing differential protein abundance between stem cell derived islet like clusters (blue circles, n=25) and donors without diabetes (teal circles, n=118). The top 40 most significant differentially abundant proteins are highlighted by labelling with gene name. **(F)** *(left)* Compares average dynamic insulin secretion stimulated by glucose (15 mM or 6 mm) or KCl (30 mM) between younger-immature (dashed brown lines, n=8) and older-maturing (solid brown lines, n=8) clusters. Basal glucose was 3 mM. Floating dot plot inserts illustrate the AUC responses for the corresponding section of the perifusion curve between younger-immature clusters (black circles) and older-maturing clusters (open circles). *(middle)* illustrates the same as *(left)* except islet-like clusters were stimulated with 5 mM leucine alone, or in combination with 6 mM glucose as depicted in figure panel. *(right)* illustrates the same as *(left)*, except islet-like were stimulated with 1.5 mM oleate/palmitate, or in combination with 6 mM glucose. **(G)** PCA plot and **(H)** volcano plot of protein abundance comparing the younger-immature clusters (light green, n=14) and older-maturing clusters (dark green, n=10).

Comparing their proteomes to human islets revealed massive differences; 3196 proteins were significantly more abundant in the non-diabetic human donor islets (Fig. 5D,E, Table S3). Some differences were due to the presence of non-islet exocrine cells trapped in our hand-picked preparations (Figs. 5E, S3). Other proteomic differences were more informative. WNT4, which we found was less abundant in lipid hyper-responders, was also more than 2 times lower in these hESC-derived islet-like clusters than in islets from non-diabetic donors (Table S3). On the other hand, 2553 proteins were more abundant in the stem cell-derived islet clusters (Table S3). Higher levels of this protein in the pancreatic beta cell are thought to limit voltage-dependent calcium channel activity, and thus decrease glucose-stimulated insulin secretion^52^. The abundance of monocarboxylate transporter 1 (SLC16A1), a beta-cell disallowed gene whose presence is suggestive a neonatal-like immature beta-cell phenotype^53^, increased over 6-fold in these islet-like clusters. Compellingly, proteins linked to important roles in fatty acid transport and metabolism long-chain fatty acid transport protein 3 (SLC27A3), mitochondrial coenzyme A transporter (SLC25A42), and solute carrier family 22 member 18 (SLC22A18) were increased ∼2-fold.

Extended maturation of stem cell-derived islet-like clusters has been shown to increase the ability of these cells to respond appropriately to a glucose stimulus^54,55^. We therefore compared nutrient-stimulated insulin secretion from younger-immature (days 21-35) and older-maturing (days 50+) stem cell preparations (Figs. 5F, S14). Nanostring analysis showed increased expression of key maturity markers (MAFA, IAPP and UCN3) in cells older than day 35 (Fig. S15). Basal insulin secretion was 5-fold higher in the older-maturing islet-like cells, and they also showed evidence of glucose-responsiveness, and maintained their sensitivity to fatty acid. At the level of the proteome, 2072 proteins more abundant in the older-maturing clusters and 2066 more abundant in the younger-immature clusters (Fig. 5G,H, Table S3). Proteins with significant roles in glucose-stimulated insulin secretion and insulin synthesis including pyruvate carboxylase (PC), mitochondrial citrate carrier (SLC25A1), high-voltage gated calcium channel subunit (CACNA2D2), glucose transporter 1 (SLC2A1), an integral SNARE protein (VAMP8) and prohormone convertase 2 (PCSK2) were all significantly more abundant in the older-maturing SC-islet clusters. These cells also had a 73% decrease in the abundance of the beta-cell disallowed gene monocarboxylate transporter (SLC16A1), when compared to more immature clusters. These proteomic data now provide a roadmap to produce more faithful human islet surrogates. Although the maturation of stem cell derived islets remains incomplete *in vitro*^55,56^, and current cell replacement tend to focus on beta-like cells, our dataset can now be used to make better stem cell-derived islets by exploiting the differences seen at the protein levels between these cells and real human islets.

## DISCUSSION

The goal of this study was to compare dynamic insulin secretion responses to carbohydrate, protein, and fat from human islets representative of the population of donors with and without type 2 diabetes, and to define the transcriptomic and proteomic mechanisms that underly the variation in response to each macronutrient. Our study is part of a large multi-institutional human islet deep phenotyping network which generates and links functional and multi-omic datasets for insight into normal and pathological variation in islet function. Our initiative provides unprecedented and unbiased mechanistic detail of the cellular processes associated with insulin secretion in response to each nutrient class, and the first to directly compare functional data to both the proteome and transcriptome. As a part of the Islet Deep Phenotyping Network Resource, this dataset forms the basis on an online platform (www.humanislets.com) providing open access to islet donor/isolation data and additional phenotyping. Despite working with high-quality research islets, all isolated from the same centre^2^, isolation parameters can have unavoidable impacts; this was particularly obvious when examining mRNA, which was more sensitive than protein to technical isolation and culture parameters. Along with weak mRNA correlation to protein in our study and others’^26,27^, this should give pause when inferring protein abundance (or function) from mRNA expression.

Our findings have relevance for personalized therapeutic nutrition and the clinical management of diabetes. For example, while both first and second phase insulin secretion in response to high glucose were blunted in islets from donors with type 2 diabetes, we did not detect significantly impaired amino acid-stimulated insulin secretion, which bolsters the case that protein-rich diets could have therapeutic benefits in patients with type 2 diabetes^57^ and highlights the need for additional research into amino-acid stimulated insulin secretion.

A recent study sought to identify differences between islets from 17 normal donors and 12 donors with type 2 diabetes, but was only able to quantify 3036 proteins, none of which were reported statistically significant after correcting for multiple comparisons^58^. Here, with more donors and more than twice the proteomic coverage, we report 555 proteins that are differentially abundant in islets from donors with type 2 diabetes. This allowed us to conduct protein-centric network analysis that was not previously possible. Proteins critical to glucose and mitochondrial metabolism coupling are less abundant in islets from donors with type 2 diabetes, explaining the blunted glucose-stimulated insulin secretory response. Conversely, proteins involved in cytoskeletal actin remodeling are on average more abundant in islets from donors with type 2 diabetes, consistent with the role of filamentous actin limiting glucose-stimulated insulin secretion in pancreatic beta cells^59^. Identifying the pathways that are perturbed in islets from donors with type 2 diabetes is crucial for better understanding the disease pathology and discovering of novel therapeutic targets.

Previous studies revealed heterogeneity in insulin secretion from human islets in response to glucose, but they do not identify the source of this variation or examine other nutrients^1–3^. Here, we leveraged the heterogeneity in nutrient-stimulated insulin secretion to identify networks of proteins that drive nutrient-specific responses. Interestingly, we identified more significant correlations between protein and insulin secretion in response to physiologically relevant 6 mM glucose, when compared to 15 mM, calling into question the routine use of such supraphysiological glucose concentrations *in vitro*.

Heterogeneity in insulin secretion in response to the three main classes of macronutrients is reminiscent of the interindividual variability of postprandial glucose responses to carbohydrates^9,60,61^. It has been suggested that this interindividual variability in glycemic response is in part responsible for the mixed weight loss outcomes following specific diet interventions. A recent randomized clinical trial did not support this idea^62^, but post-prandial glucose and insulin responses were not measured to confirm the effectiveness of the reported personal diets. On the other hand, a diet specifically targeting hyperinsulinemia was shown to be highly effect in the prevention of multiple diseases^7^. Multi-omic profiling of blood plasma recently revealed heterogeneity in individual responses to a “healthy lifestyle intervention”^63^. Combined with our results, these studies provide the rationale for a clinical trial to test the insulin response to standardized macronutrients challenges in the general population of normoglycemic individuals and those living with type 2 diabetes.

One of our most surprising findings was that ∼8% of donor islets showed relative hyperresponsiveness to a fat stimulus at basal glucose. Strikingly, the greatest number of correlations to protein abundance were in response to the fatty acid stimulus, meaning that these islets are quite different from those exhibiting the typical response ratio. We speculate that this lipid responsiveness is related to beta cell immaturity because it is also observed in stem-cell derived immature cells. The magnitude of fatty acid-stimulated insulin secretion has been debated^4,14,64^, and we are not aware of clinical literature that has reported on individuals with higher lipid-stimulated insulin secretion compared with glucose-stimulated insulin secretion *in vivo*. Studies with sufficient power and participant diversity have not been conducted and reported yet. Our literature search of clinical studies examining the effects of glucose and oral or iv lipid infusions in human participants was only able to find studies with low n numbers that did not provide detailed information on the response distribution (Table S11)^65–70^, so direct comparisons with our dataset are not possible. We hope our results will encourage clinical researchers to look for response heterogeneity for each macronutrient in heterogenous cohorts >100.

In summary, we present evidence that nutrient-specific insulin secretion is heterogeneous in the general population. In the face of hyperinsulinemia being causal to numerous health conditions, our benchmark study lays the groundwork for future clinical studies aiming to advance the area of personalized therapeutic nutrition.

## DATA AND MATERIALS AVAILABILITY

All data are available, both via a purpose-build web portal and the appropriate public databases. The raw RNA sequencing data has been deposited in the European Genome-phenome Archive (EGA) under study number EGAS00001007241. The raw proteomics data has been deposited to ProteomExchange via MASSive under dataset number PXD045422. This dataset, together with the functional data will also be available for download and browsing on a new portal website www.humanislets.com.

## MATERIALS AND METHODS

### Human islet culture

Islets were isolated from pancreas of cadaveric human donors at the Alberta Diabetes Institute IsletCore and used with approval of the Human Research Ethics Board at the University of Alberta (Pro00013094; Pro00001754), the University of British Columbia (UBC) Clinical Research Ethics Board (H13-01865), the University of Oxford’s Oxford Tropical Research Ethics Committee (OxTREC Ref.2-15), the Oxfordshire Regional Ethics Committee B (REC reference: 09/H0605/2), or by the Stanford Center for Biomedical Ethics (IRB Protocol: 57310). T2D diagnosis was family-declared at the time of organ donation. All families of organ donors provided written informed consent for use in research. Islet isolation was performed according to the detailed methods deposited in the protocols.io repositor ^71^. Islets were shipped in CMRL media (Thermo Fisher Scientific) overnight from Edmonton to UBC. Upon arrival, islets were immediately purified by handpicking under a stereomicroscope and suspended in RPMI 1640 medium (Thermo Fisher Scientific, Cat#, 11879-020) supplemented with 5.5 mmol glucose, 10% fetal bovine serum (FBS), and 100 units/mL penicillin/streptomycin. Islets were cultured in 10 cm non-adhesive petri dishes (Thermo Fisher Scientific, Cat # FB0875713) at a density of 10-16 islets/cm^2^ (100-160 IEQ/ cm^2^) for 24-72 hours prior to experiments to allow for recovery from shipment. Total islet culture time (as well as additional donor and isolation characteristics) are listed in Table 1.

### Deep dynamic phenotyping of human and mouse islets, and stem cell-derived islet-like clusters

Our standard approach ^72^ compared the response to 6 mM or 15 mM glucose stimulation and direct depolarization with 30 mM KCl. In parallel, we also measured insulin secretion in response to 5 mM leucine (Sigma, Cat. # L8912, dissolved in 1M HCl, then pH adjusted with 1M NaOH) – an essential branched chain amino acid previously shown to increase insulin secretion in human islets^5^; or a 1:1 mixture of 0.75 mM oleic acid (Sigma, Cat. # 364525) and 0.75 mM palmitic acid (Sigma, Cat. #P5585) at either 3 mM or 6 mM glucose (see figure legends). Fatty acids stock solutions were prepared in 50% ethanol at 65°C for 30 minutes, and then added to a solution of fatty acid free bovine serum albumin (BSA) (Sigma, Cat. # A7030) in a 6:1 molar ratio in a 37 °C water bath for 60 min.

We loaded perifusion columns with either 65 human islets, 100 (hESC)-stem cell-derived islet-like clusters, or 100 mouse islets and perifused them at 0.4 mL/min with 3 mM glucose Krebs-Ringer Modified Buffer (KRB) solution as described previously ^72^ for 60 minutes to equilibrate the islets to the KRB and flow rate, and then with the indicated conditions. First-phase insulin release was defined as the amount of insulin secreted during the first 15 minutes of 15 mM glucose stimulation, while the remaining 25 min of stimulation were defined as second-phase release. Peak insulin secretion was defined as single point whereby the amount of insulin released during the first 15 minutes of a solution change was the highest. Samples were stored at –20 °C and insulin secretion was quantified using human (Millipore Cat. # HI-14K) or rat (Millipore Cat. # RI-13K) insulin radioimmunoassay kits. All insulin secretion units have been converted to pmol/islet, to allow for direct comparison between species.

### Mouse islet isolation and culture

C57BL/6J mice were purchased from the Jackson Laboratory and housed in the UBC Modified Barrier Facility (temperature-controlled) using protocols approved by the UBC Animal Care Committee and in accordance with international guidelines. The mice were on a 12-hour light/dark cycle with chow diet (LabDiet #5053) and drinking water *ad libitum*. Mouse islets were isolated by ductal collagenase (Type XI; Sigma, Cat. # C7657) injection followed by hand-picking. To allow for recovery post digestion, islets were cultured overnight in islet culture media (RPMI media with 11.1 mM D-glucose supplemented with 10% vol/vol fetal bovine serum (Thermo Fisher Scientific, Cat. #12483020) and 1% vol/vol Penicillin-Streptomycin (Gibco, Cat #15140-148)) at 37°C with 5% CO_2_.

### Human embryonic stem cell-derived islet-like cluster culture and differentiation

A human embryonic stem cell line, based on the WiCell WA01 line, that contained knock-in add-on of EGFP downstream of the insulin coding sequence ^51^ was differentiated, as previously described, through a 6-stage protocol to day 21, media components are included in the table below^51,73^. After the initial 21 days of differentiation, SC-β cells were cultured in CMRL with 5.6 mM glucose (Thermo Fisher Scientific) containing 1% fatty acid-free BSA (Proliant), 1:100 Glutamax, 1:100 NEAA, 1 mM pyruvate, 10 mM HEPES, 1:100 ITS (Thermo Fisher Scientific), 10 μg/ml of heparin sulfate, 1 mM N-acetyl cysteine, 10 μM zinc sulfate, 1.75 μL 2-mercaptoethanol, 2 μM T3 and 0.155 mM ascorbic acid (Sigma Aldrich) until Day 27. Between Day 28-50, aggregates were grown in CMRL with 5.6 mM glucose containing 1% fatty acid-free BSA, 1:100 Glutamax, 1:100 NEAA, 1 mM Pyruvate, 10 mM HEPES, 1:100 ITS, 10 μg/ml heparin sulfate, 1 mM N-acetyl cysteine, 10 μM zinc sulfate, 1.75 μL 2-mercaptoethanol, 10 nM T3, 1:2000 Trace elements A (Cellgro), 1:2000 trace elements B (Cellgro), 1:2000 Lipid Concentrate (Thermo Fisher Scientific) and 0.5 µM ZM447439 (Selleckchem) ^55^.

Dispersion/reaggregation of the stem cell-derived aggregates was carried out on Day 25 as previously described ^74^. Briefly, aggregates were dissociated with Accumax (Thermo Fisher Scientific) for 8 minutes at 37 **°**C with occasional shaking. Cells were washed with DPBS, centrifuged (5min/200xg) then resuspended in DPBS with 0.2% FBS and 10µM Y-27632. Cells were filtered through a strainer-capped FACS tube prior to FACS purification of the GFP+ fraction. FAC-sorted cells were collected in Stage 6.2 media containing 10µM Y-27632 and then aggregated in AggreWell 400 24-well plates (STEMCELL Technologies) using the manufacturer’s protocol at 1000 cells per aggregate.

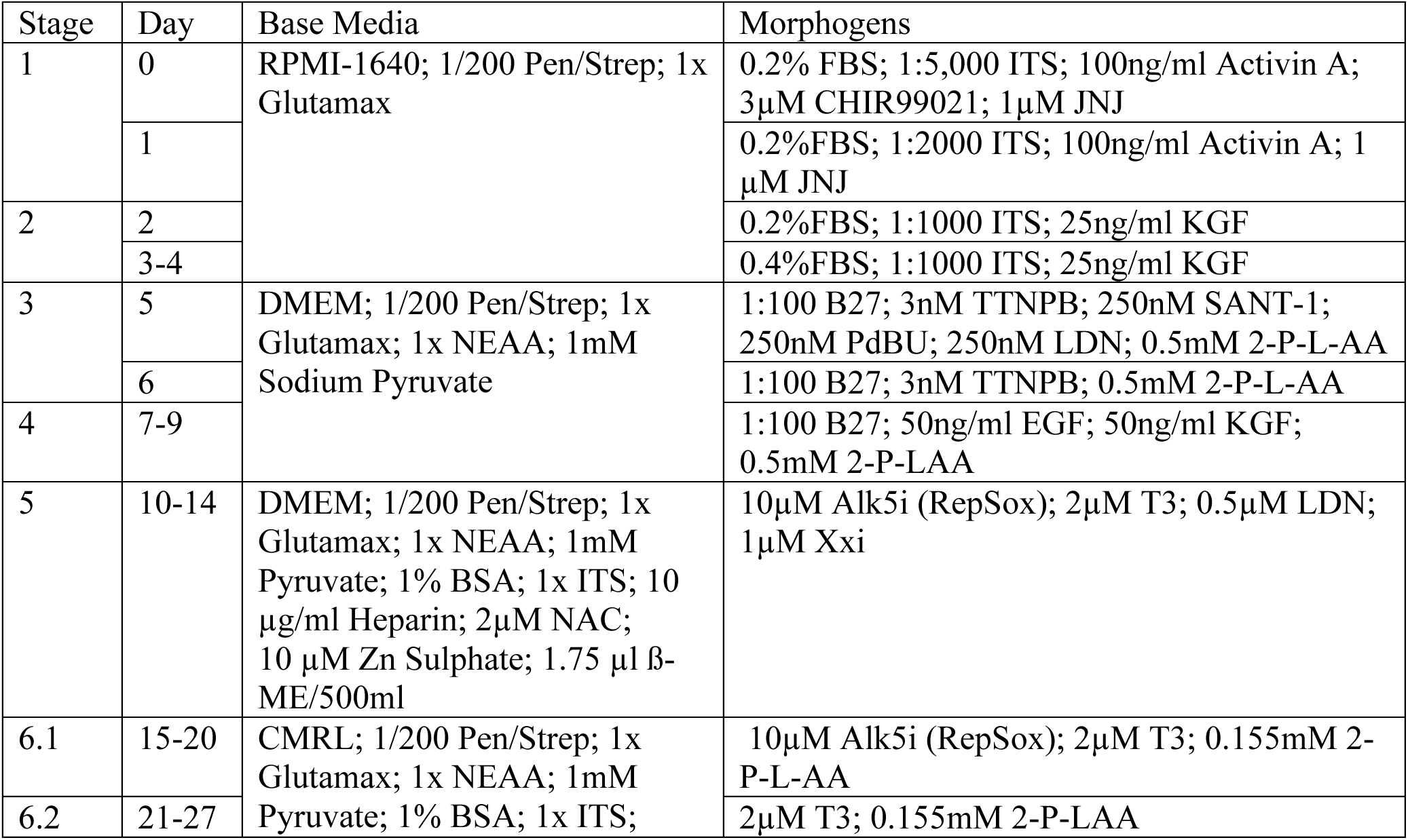
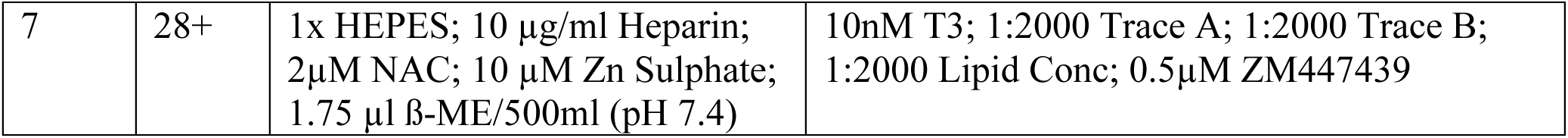
Media formulations.

### Proteomics

#### Library generation

We generated a pooled protein sample of 10 non-diabetic donors (as indicated in Table S1) at a concentration of 1.5ug/uL. Briefly, 100 μl of SDS lysis buffer (4 % SDS, 100 mM Tris, pH = 8) was added to each sample. Samples were then vortexed for 1 minute, boiled at 100°C for 10 minutes, vortexed again for 1 minute and then centrifuged at 4°C at 10,000 RPM for 10 minutes. The supernatants were collected and tested for protein concentration by BCA protein assay (ThermoFisher Scientific, Cat. # PI-23225). Measurements were performed on a Spark plate reader (TECAN). All protein samples were stored at –80°C.

Next, 297 μg of the pooled protein sample was used for further processing. The Trapped Ion Mobility – Time of Flight Mass Spectrometer (TimsTOF Pro; Bruker Daltonics, Germany) was set to Parallel Accumulation-Serial Fragmentation (PASEF) scan mode for DDA acquisition scanning 100 – 1700 m/z with 5 PASEF ramps. The capillary voltage was set to 1800V, drying gas to 3L/min, and drying temperature to 180°C. The MS and MS/MS spectra were acquired from m/z 100 – 1700. As for TIMS setting, ion mobility range (1/k0) was set to 0.70 – 1.35 V·s/cm^2^, 100ms ramp time and accumulation time (100% duty cycle), and ramp rate of 9.42Hz; this resulted in 0.64s of total cycle time.

Linear precursor repetitions were applied with a target intensity of 21,000 and 2500 intensity threshold. The active exclusion was enabled with a 0.4 min release. The collision energy was ramped linearly as a function of mobility from 27eV at 1/k0 = 0.7 V·s/cm^2^ to 55eV at 1/k0 = 1.35 V·s/cm^2^. Isolation widths were set at 2.07 m/z at < 400 m/z and 3.46 m/z at > 1000 m/z. Mass accuracy: error of mass measurement is typically within 3 ppm and is not allowed to exceed 7 ppm. For calibration of ion mobility dimension, the ions of Agilent ESI-Low Tuning Mix ions were selected (m/z [Th], 1/k0 [Th]: 622.0290, 0.9915; 922.0098, 1.1986; 1221.9906, 1.3934). TimsTOF Pro was run with timsControl v. 3.0.0 (Bruker). LC and MS were controlled with HyStar 6.0 (6.0.30.0, Bruker).

Acquired data were then searched using FragPipe^75^ computational platform (v. 17.1) with MSFragger^76,77^ (v. 3.4), Philosopher^78^ (v. 3.8), and EasyPQP (v. 0.1.27) components to build a spectral library. Protein sequence database Homo sapiens from Uniprot (reviewed sequences only; downloaded on January 15, 2021) and common contaminants protein, containing in total 20428 sequences were used, where reversed protein sequences were appended to the original database as decoys. For the MSFragger analysis, both precursor and fragment mass tolerances were set to 50 ppm. Enzyme specificity was set to “trypsin”, with up to 2 missed cleavages allowed.

For each analysis, the MS/MS search results were further processed using Philosopher, where final reports were generated and filtered at 1% protein FDR plus 1% PSM/ion/peptide-level FDR^79^ for each corresponding PSM.tsv, ion.tsv, and peptide.tsv files. Finally, .tsv files were used as input to EasyPQP for the generation of consensus spectrum libraries. The final spectral library was filtered to 1% protein and 1% peptide-level FDR.

#### Experimental samples

For each donor approximately 300 hand-picked islets were transferred into a 1.5 ml microcentrifuge tube. Culture media was removed, islets were washed twice in 1X PBS (Invitrogen, Cat. #10010049) and cell pellets were immediately flash frozen in liquid nitrogen and then stored at –80°C. Samples were processed and protein was extracted and measured as described above.

Next, 20 μg of each lysed sample was taken for further processing. Reduction of disulfide bonds was done by incubation with DTT (30 mM) for 35 min at 37°C, followed by alkylation by CAA (50 mM) for 20 min at 37°C. Samples were loaded onto 10% Mini-PROTEAN® TGX™ Precast Protein Gels (BioRad) and run for 30 min at 85V. Proteins were visualized by Coomassie blue (stained for 20 min). Lanes were cut out and destained in Ambic:EtOH (60:40), dehydrated and digested with Trypsin, first round 0.32ug/lane for 15 hours at 37°C, second round 0.128ug/lane for 2 hours, at 37°C (0.448ug in total/lane). Digestion was stopped with 10% FA, and samples were extracted with a series of extraction solutions, twice by 50% ACN, 50% of 0.1% TFA, and twice by 80% ACN, 20% of 0.1% TFA. Samples were then concentrated via vacuum centrifugation. Extracted peptide samples were then cleaned up via STAGE-tip purification, briefly: Resolubilized acidified sample was forced through a conditioned and equilibrated column with 8-12 mm of C18 packing, washed with 1% TFA twice, and eluted into clean plates by buffer containing 40% ACN, 0.1% TFA, then dried down.

Each sample was reconstituted in 0.5% ACN, 0.1% formic acid for LC-MS/MS analysis of 150 ng total on-column injections (with n = 3 technical replicates). The digest was separated using NanoElute UHPLC system (Bruker Daltonics) with Aurora Series Gen2 (CSI) analytical column (25 cm x 75 μm 1.6 μm FSC C18, with Gen2 nanoZero and CSI fitting; Ion Opticks, Parkville, Victoria, Australia) heated to 50°C and coupled to a Trapped Ion Mobility – Time of Flight mass spectrometer (timsTOF Pro; Bruker Daltonics, Germany) operated in Data-Independent Acquisition – Parallel Accumulation-Serial Fragmentation (DIA-PASEF) mode. A standard 60 min gradient was run from 2% B to 12% B over 30 min, then to 33% B from 30 to 60 min, then to 95% B over 0.5 min, and held at 95% B for 7.72 min. Before each run, the analytical column was conditioned with 4 column volumes of buffer A. Where buffer A consisted of 0.1% aqueous formic acid and 0.5 % acetonitrile in water, and buffer B consisted of 0.1% formic acid in 99.4 % acetonitrile. The NanoElute thermostat temperature was set at 7°C. The analysis was performed at 0.3 μL/min flow rate.

The TimsTOF Pro was set to Parallel Accumulation-Serial Fragmentation (PASEF) scan mode for DIA acquisition scanning 100 – 1700 m/z. The capillary voltage was set to 1800V, drying gas to 3L/min, and drying temperature to 180°C. The MS1 scan was followed by 17 consecutive PASEF ramps containing 22 non-overlapping 35 m/z isolation windows, covering the m/z range 319.5 – 1089.5 (more information in DIA windows). As for TIMS setting, ion mobility range (1/k0) was set to 0.70 – 1.35 V·s/cm^2^, 100ms ramp time and accumulation time (100% duty cycle), and ramp rate of 9.42 Hz; this resulted in 1.91s of total cycle time. The collision energy was ramped linearly as a function of mobility from 27eV at 1/k0 = 0.7 V·s/cm^2^ to 55eV at 1/k0 = 1.35 V·s/cm^2^. Mass accuracy: error of mass measurement is typically within 3 ppm and is not allowed to exceed 7 ppm. For calibration of ion mobility dimension, the ions of Agilent ESI-Low Tuning Mix ions were selected (m/z [Th], 1/k0 [Th]: 622.0290, 0.9915; 922.0098, 1.1986; 1221.9906, 1.3934). TimsTOF Pro was run with timsControl v. 3.0.0 (Bruker). LC and MS were controlled with HyStar 6.0 (6.0.30.0, Bruker).

Acquired diaPASEF data were then searched using FragPipe ^78^ computational platform (v. 17.1) with MSFragger ^79,80^ (v. 3.4), Philosopher ^78^ (v. 3.8), EasyPQP (v. 0.1.27) and DIA-NN (v. 1.8) to obtain DIA quantification, with use of the spectral library generated from the before mentioned highly fractionated sample. Quantification mode was set to “Any LC (high precision)”. All other settings were left default.

### RNAseq and Nanostring

Islets for RNA-seq were processed in two centers, initially in Oxford (n=49) with later donors being processed at Stanford (n = 47). For those processed in Oxford the methods have been described ^81^. Briefly, freshly isolated human islets were collected at the Alberta Diabetes Institute IsletCore (www.isletcore.ca) in Edmonton, Canada. Freshly isolated islets were processed for RNA and DNA extraction after 1-3 days in culture in CMRL media. RNA was extracted from human islets using Trizol (Ambion, UK or Sigma-Aldrich, Canada). To clean remaining media from the islets, samples were washed three times with phosphate-buffered saline (Sigma-Aldrich, UK). After the final cleaning step 1 mL Trizol was added to the cells. The cells were lysed by pipetting immediately to ensure rapid inhibition of RNase activity and incubated at room temperature for 10 min. Lysates were then transferred to clean 1.5 mL RNase-free centrifuge tubes (Applied Biosystems, UK). RNA quality (RIN score) was determined using an Agilent 2100 Bioanalyser (Agilent, UK), with a RIN score > 6 deemed acceptable for inclusion in the study. Samples were stored at −80 °C prior to sequencing. PolyA selected libraries were prepared from total RNA at the Oxford Genomics Centre using NEBNext ultra directional RNA library prep kit for Illumina with custom 8 bp indexes ^82^. Libraries were multiplexed (three samples per lane), clustered using TruSeq PE Cluster Kit v3, and paired-end sequenced (75bp) using Illumina TruSeq v3 chemistry on the Illumina HiSeq2000 platform. For the samples processed at Stanford, freshly isolated human islets were collected at the Alberta Diabetes Institute IsletCore in Edmonton, Canada, the same as the Oxford samples. The islets were picked to >95% purity and washed with PBS, then stored in 1 mL of Trizol (Ambion, UK). Islets in trizol were stored at –80 C until they were processed for RNA using the phenol-chloroform extraction method. The extracted RNA was resuspended in water and then dnased to remove contaminating DNA. RNA quality (RIN score) and concentration were determined using an Agilent 2100 Bioanalyzer (Agilent, US), with a RIN score > 5 deemed acceptable for inclusion in the study. Samples were stored at −80 °C prior to sequencing. PolyA selected cDNA libraries were prepared from total RNA and then sequenced using NovaSeq 6000 by Novogene. The raw sequencing data (fastq files) were checked first in regarding of the number of reads (>= 20 million per sample and extra sequencing were performed if necessary) and then in regarding of quality control, including quality score per base and length distribution using fastqc (v0.11.9). Reads were aligned to human genome reference (GRCh38) using STAR (v2.7.9a) with ENSEMBL gene annotations (v101). Gene expression levels were counted using featureCounts (v2.0.1) on exonic reads only. Differential expression was compared using the Wald test in DESeq2 (v1.26.0). *p* values were adjusted using the Benjamini and Hochberg method.

Fifty islets from each donor batch were used to assess islet quality by profiling expression of 132 human islet genes as described ^83^. Gene expression was measured using nCounter prep kits and nCounter SPRINT profiler according to manufacturer’s protocol (NanoString, USA).

### Co-expression analysis

Analysis was performed with both RNA-seq and proteomics data to assess relationships between ‘modules’ of omics features and donor metadata, including technical islet isolation parameters, clinical metadata, and functional outcomes. There were three main steps: data processing, network construction and functional characterization, and module-donor metadata correlation analysis.

Raw protein abundance matrices (134 donors, 8489 proteins; Uniprot IDs) were filtered to remove proteins with greater than 50% missing values, reducing the number of proteins to 7919. Next, the proteomics data were normalized by sample median, log transformed (base 10), and missing values were imputed with the *missForest* R package ^84^ using a random forest-based algorithm. The RNA-seq counts matrix (96 donors, 17 673 genes; Entrez IDs) was filtered based on abundance (15% with lowest mean counts removed), normalized with the ‘variance stabilizing transformation’ from the *DeSeq2* R package ^85^, and then filtered based on variance (15% with lowest variance removed). The RNA-seq data was measured in seven batches; batch effects were adjusted for using the ComBat method in the *sva* R package ^86^. One of the batches (*n* = 6 samples) had a sequencing depth of about 50% compared to the other samples, resulting in ∼5000 genes with zero counts that prevented effective batch effect adjustment. These six samples were removed from the dataset. Donor metadata included clinical metadata (age, sex, BMI, diabetes diagnosis, HbA1c), technical islet isolation metadata (culture time, cold ischemia time, digestion time, purity, and insulin content), and functional outcomes (AUC values from perifusion experiments after glucose, leucine, and oleate/palmitate exposures). Continuous variables with right-skew distributions were log-transformed (base 10). A constant value was added to the perifusion outcomes that had negative values to ensure all data were > 0 before log-transformation.

Two co-expression networks were constructed using the same methods, one for each omics type, using the *WGCNA* R package ^87^. First, the soft threshold parameter was selected by choosing the lowest value at which the R^2^ value of the scale free topology model fit did not substantially increase (threshold parameter = 8 and 20 for proteomics and RNA-seq respectively; selected via visual examination of an elbow plot). Next, pairwise distances between omics features were calculated by computing signed adjacency and topological overlap matrices using the soft threshold parameter and the ‘bicor’ correlation function. Finally, modules of co-expressed (or co-abundant) features were defined by first hierarchically clustering features based on the distance matrix, and then detecting modules of densely connected features using the dynamic tree cut algorithm. Modules were annotated with KEGG pathways and GO terms (biological process, cellular component, and molecular function) by performing overrepresentation analysis with the list of features in each module.

The pairwise partial correlation between module expression/abundance levels and donor metadata, accounting for the technical isolation parameters (purity, culture time, digestion time, and cold ischemia time), were calculated with the partial Pearson correlation function in the *ppcor* R package ^88^. Module expression/abundance levels were summarized with *WGCNA* ‘eigengenes’, which are the first principal component scores for each module, re-scaled so that positive and negative scores can be interpreted as higher and lower expression/abundance levels respectively. Correlation p-values were adjusted using the Benjamini-Hochberg method, with the FDR set to 5%.

### Multi-omics data analyses

RNAseq data was analyzed by DESeq2 R package (version 1.36.0)^85^. Six samples exhibiting large batch difference were excluded, leaving a total of 90 individuals (82 ND and 8 T2D) included in the analyses. Genes with low counts (<5 raw counts in more than 50% samples) were removed. Raw counts of the genes were normalized by variance stabilizing transformation (VST). T2D group was contrasted to ND group, including batch as a covariate in the default Wald test. Genes with Benjamini-Hochberg adjusted p-values < 0.05 were considered as differentially expressed.

For proteomics data analyses, proteins that were undetected in more than 50% samples were removed. Benjamini-Hochberg adjusted p-values < 0.05 of student t-test were used to identify differentially expressed proteins between the various identified groups.

To identify proteins associated with islet parameters, linear regression was conducted using log2 protein abundances of each protein as a dependent variable, each islet parameter as an independent variable, and diabetes status as a covariate to adjust for the effect of T2D. The coefficients of islet parameters and their FDR-adjusted p-values were reported. Similarly, to identify RNAs associated with islet parameters, the VST normalized gene counts were used as dependent variables instead. The batches of sequencing were included as additional covariates. Pearson correlations were also conducted instead of linear regression.

To assess the how well RNA expression reflects the abundance of the corresponding protein, we conducted Pearson correlation between proteins and RNAs. For across-gene correlation, mean protein abundances and mean RNA TPMs (transcript per million) were correlated. We used protein signal/protein length as a raw estimate of relative abundances across different proteins and TPM as an estimate of relative RNA levels across different genes. For within-gene across sample correlations, we correlated each gene’s VST normalized RNA counts with log2 protein abundances, and the p values were adjusted by Benjamini-Hochberg. Instead of Pearson correlation, we also checked within-gene association using linear regression adjusting for batches of sequencing, and the results were mostly the same (not shown).

Gene set enrichment analysis (GSEA) was performed on the transcriptomic or proteomics changes (T2D/ND) using KEGG or Hallmark gene set libraries. The direction-signed –log_10_(p values) were used as rank scores. Over representation analysis (ORA) was performed on genes with positive, negative or non-significant RNA-protein correlations using Gene Ontology – Cellular Component (GO-CC) gene set library. Genes that are commonly detected in RNAseq and proteomics were used as background gene list. Both GSEA and ORA were conducted in clusterProfiler R package^89^.

To evaluate the concordant changes of RNAs and proteins in T2D, in addition to comparing differentially expressed RNAs and proteins, we applied the threshold-free method called rank-rank hypergeometric overlap test^19^. RNAs and proteins were ranked by direction-signed –log10(p value) or log2 fold change (T2D/ND). The ranked lists were processed in the online tool (https://systems.crump.ucla.edu/rankrank/) and RRHO R package using a step size of 100.

Cell type deconvolution was conducted using the BisqueRNA R package using a marker-based decomposition approach. RNAseq DESeq2 normalized counts (without VST) or protein abundances (without log2 transformation) were used as the input. Islet cell type markers (alpha, beta, gamma, delta) were from van Gurp et al. ^90^, and acinar cell markers were from Segerstolpe ^91^. All code can be found at Github repository https://github.com/hcen/Kolic_macronutrients.

### General statistical analyses and data visualization

Statistical analyses and data presentation for perifusion studies of dynamic insulin secretion were carried out using GraphPad Prism 9 (Graphpad Software, San Diego, CA, USA) or R (v 4.1.1). using Student’s *t*-test for parametric data and Mann-Whitney test for non-parametric data. For all statistical analyses, differences were considered significant if the p-value was less than 0.05. *: p< 0.05; ** p< 0.01; *** p< 0.001. Data were presented as means ± SEM with individual data points from biological replicates (average of two technical replicates).

## AUTHOR CONTRIBUTIONS

Conceptualization: JK, JX, FCL, ALG, LJF, PEM, JDJ

Methodology: JK, HHC, JE, HS, VR, SS, AFG, JEM, JL, LB, JX, FCL, ALG, LJF, PEM, JDJ

Investigation: JK, RM, JCR, HS, SS, FCL, ALG

Visualization: JK, WGS, HC, JE, YHX, JDJ

Funding acquisition: JX, FCL, ALG, LJF, PEM, JDJ

Supervision: JX, FCL, ALG, LJF, PEM, JDJ

Writing – original draft: JK, JDJ

## Data Availability

All data produced in the present study are available upon reasonable request to the authors.

## ACKNOWLEDGMENTS

We thank the Human Organ Procurement and Exchange (HOPE) program and Trillium Gift of Life Network (TGLN) for their work procuring human donor pancreas for research. We especially thank the organ donors and their families for their gift in support of diabetes research. The authors acknowledge that work done at UBC and BC Children’s Hospital occurs on the traditional, ancestral, and unceded territories of the Coast Salish peoples, the Sḵwx̱ wú7mesh (Squamish), sə^l̓^ ilwətaɁɬ (Tsleil-Waututh), and xʷməθkʷə^y̓^ əm (Musqueam) Nations.

