## Supplemental Figures for "Proteomic predictors of individualized nutrient-specific insulin secretion in health and disease"

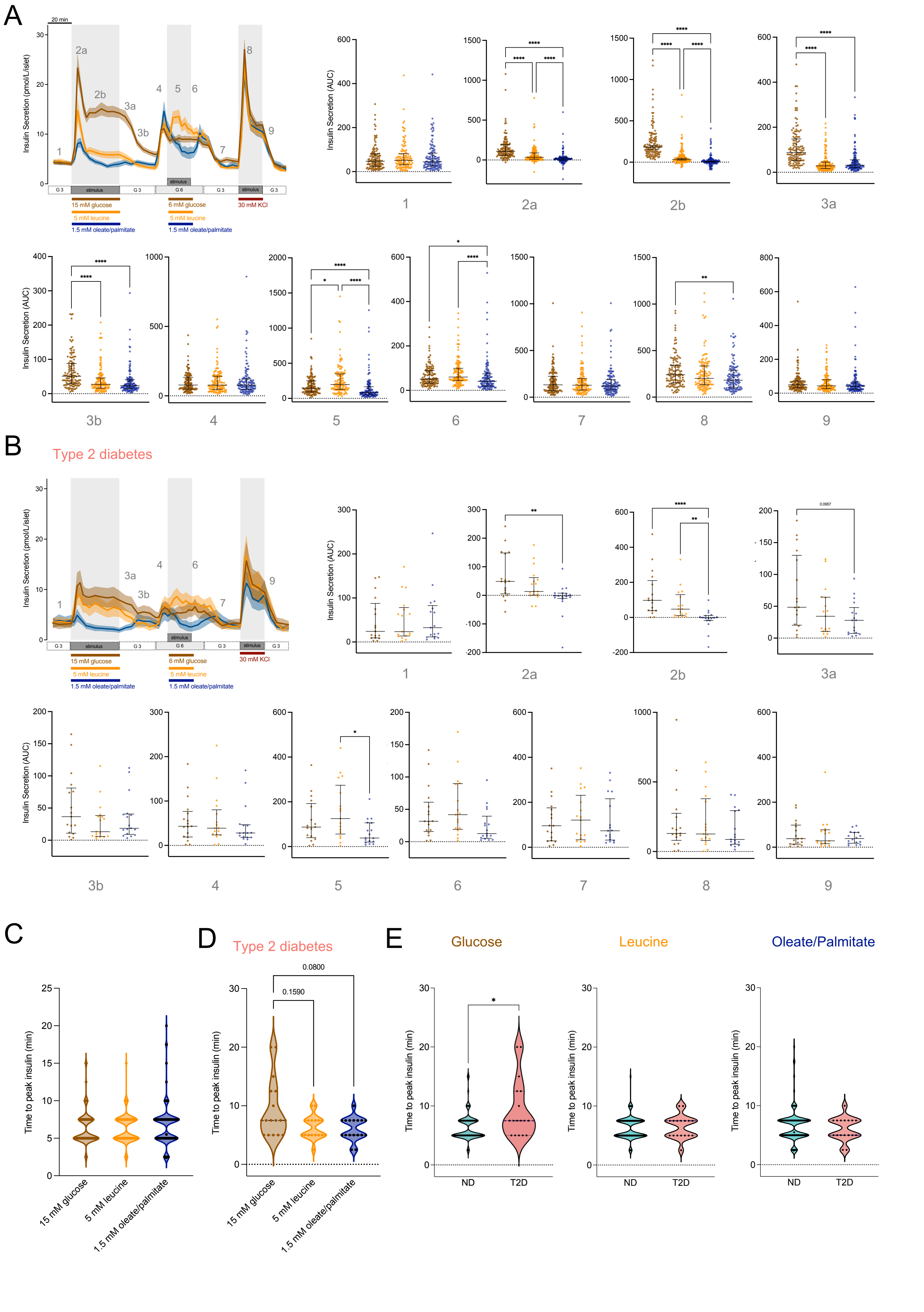
A
B
Type 2 diabetes
C
D
Type 2 diabetes
E
Glucose
Leucine
Oleate/Palmitate
Insulin Secretion (pmol/L/islet)
Insulin Secretion (AUC)
Time to peak insulin (min)
20 min
15 mM glucose
5 mM leucine
1.5 mM oleate/palmitate
6 mM glucose
30 mM KCl
stimulus
G 3
G 6
15 mM glucose
5 mM leucine
1.5 mM oleate/palmitate
ND
T2D
0.1590
0.0800
0.0957

Figure S1

**Fig. S1. Average dynamic insulin secretion perifusion AUC calculations in non-diabetic donors and donors with type-2 diabetes. (A)** Averaged traces of dynamic insulin secretion measurements in response to glucose (15 or 6 mM), leucine (5 mM), 1.5 mM oleate and palmitate (1:1 mixture) or 30 mM KCl in islets isolated from non-diabetic donors (n=123). Numbers in grey represent the following time intervals: 1. baseline 3mM glucose (0-20 min) 2a. 1st phase (20-35 min) 2b. second phase (35-60 min) 3a. 3mM glucose (60-75 min) 3b. 3mM glucose (75-90 min) 4. 6 mM glucose (90-100 min) 5. 6 mM glucose (100-120 min) 6. 6 mM glucose (120-130 min) 7. 3mM glucose (130-160 min) 8. 30 mM KCl (160-180 min) and 9. 3 mM glucose (190-200 min). Corresponding areas under the curve for each indicated section, with median and interquartile range are shown. **(B)** Shows the same as (A) except in islets from donors previously diagnosed with type 2 diabetes (n=17). **(C)** Time-to-reach peak insulin secretion following a 15 mM glucose stimulus (brown), 5 mM leucine stimulus (orange) or 1.5 mM oleate/palmitate stimulus (blue) in non-diabetic donors (n=123). **(D)** Shows the same as (C) except in islets from donors diagnosed with type 2 diabetes (n=17). **(E)** Shows the direct comparisons of time-to-reach peak between non-diabetic donors (n=123) and donors with type-2 diabetes (n=17) for each macronutrient tested.

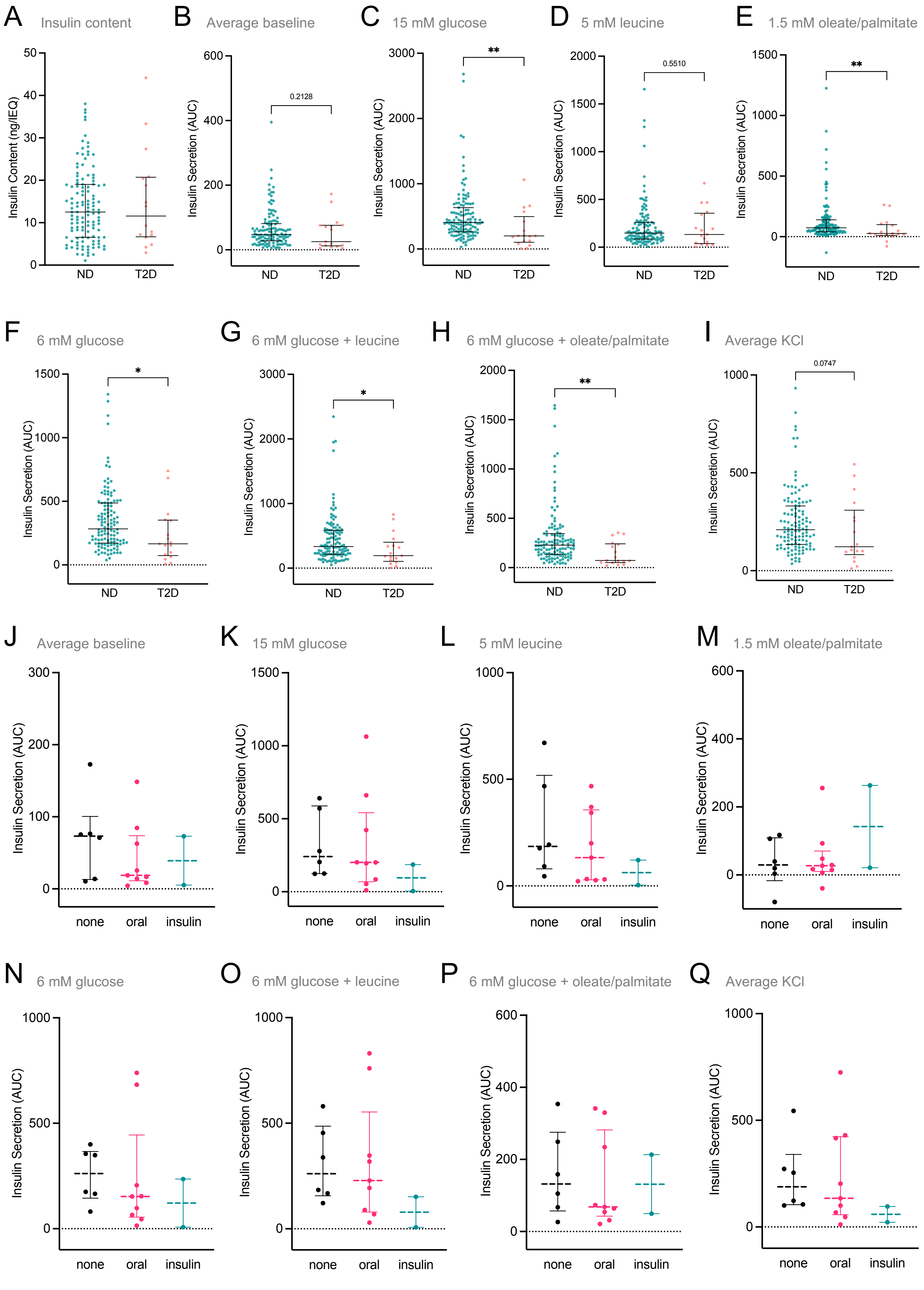

A Insulin content
Insulin Content (ng/IEQ)
ND
T2D
B Average baseline
0.2128
C 15 mM glucose
**
D 5 mM leucine
0.5510
E 1.5 mM oleate/palmitate
**
F 6 mM glucose
*
G 6 mM glucose + leucine
*
H 6 mM glucose + oleate/palmitate
**
I Average KCl
0.0747
J Average baseline
K 15 mM glucose
L 5 mM leucine
M 1.5 mM oleate/palmitate
N 6 mM glucose
O 6 mM glucose + leucine
P 6 mM glucose + oleate/palmitate
Q Average KCl
Insulin Secretion (AUC)
none
oral
insulin


Figure S2

**Fig. S2. Comparing nutrient-stimulated insulin secretion in non-diabetic donors and donors with type-2 diabetes. (A)** Islet insulin content (ng/IEQ) comparison between non-diabetic donors and donors with type-2 diabetes. Median and interquartile range are shown. **(B-I)** Direct comparisons of AUC values between non-diabetic donors and donors with type-2 diabetes for indicated areas of the perifusion curve. Median and interquartile range are shown. **(J-Q)** Shows direct comparisons of AUC values of donors with type-2 diabetes grouped by known medication status. “None” indicates that the donors were not taking any known glucose-lowering medications. “Oral” indicates that donors were taking oral hypoglycemic mediations (metformin in most cases). “Insulin” indicates that the donors have progressed to needing exogenous insulin for glucose control. Median and interquartile range are shown.

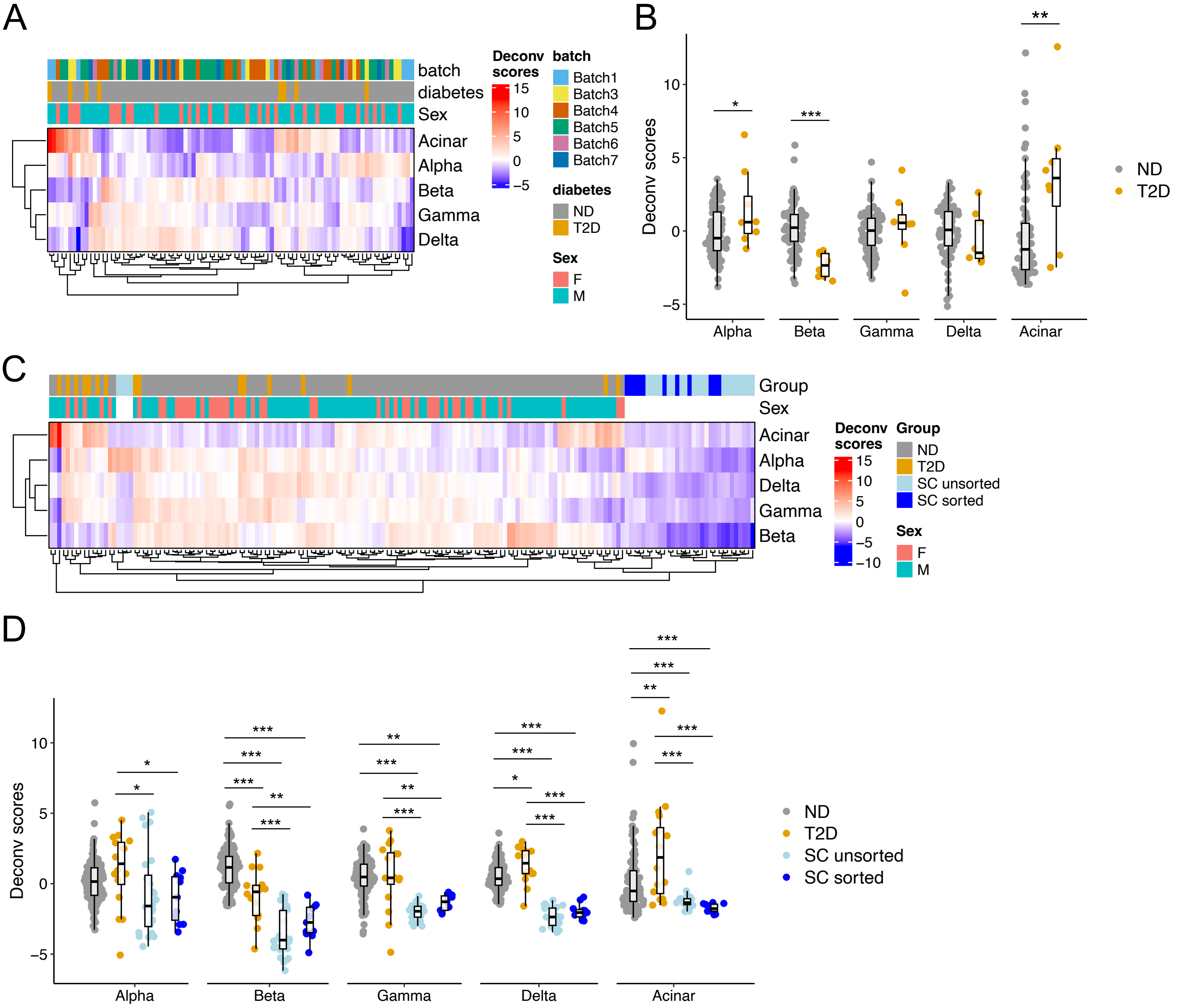


**Fig. S3. Islet cell type deconvolution scores of human islets and stem cell-derived islet clusters from RNAseq or proteomics. (A)** Heatmap showing the deconvolution scores from human islet RNAseq. Batches, diabetes status and sex are annotated on top. **(B)** Deconvolution scores from human islet RNAseq comparing ND and T2D donors. **(C)** Heatmap showing the deconvolution scores from human islet or stem cell-derived islet cluster proteomics. **(D)** Deconvolution scores from proteomics comparing ND and T2D human islets and unsorted or sorted stem cells (SC)-derived islet clusters.

Figure S3

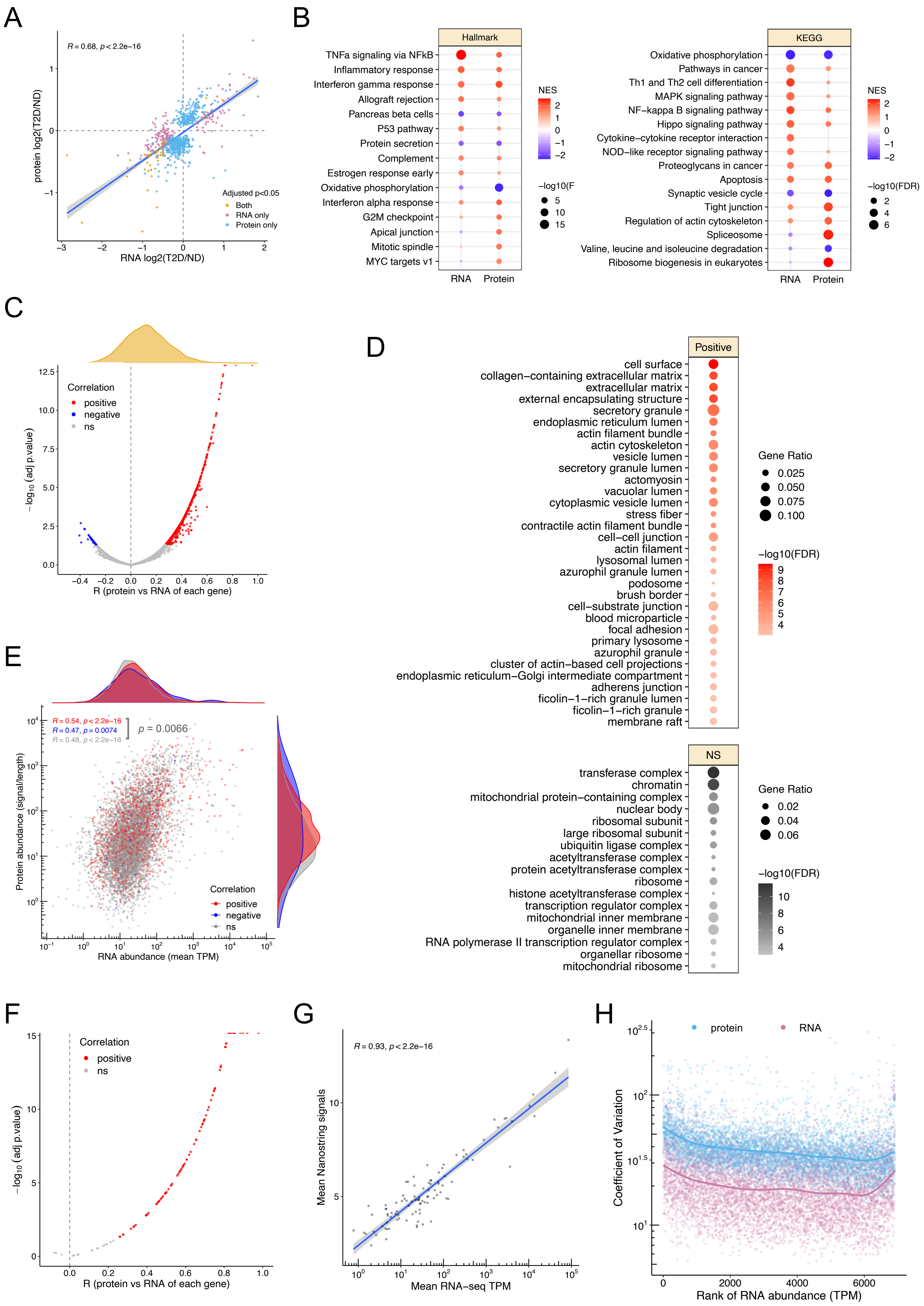

A
R = 0.68, p < 2.2e−16
protein log2(T2D/ND)
RNA log2(T2D/ND)
Adjusted p<0.05
Both
RNA only
Protein only
B
Hallmark
TNFa signaling via NFkB
Inflammatory response
Interferon gamma response
Allograft rejection
Pancreas beta cells
P53 pathway
Protein secretion
Complement
Estrogen response early
Oxidative phosphorylation
Interferon alpha response
G2M checkpoint
Apical junction
Mitotic spindle
MYC targets v1
RNA
Protein
NES
KEGG
Oxidative phosphorylation
Pathways in cancer
Th1 and Th2 cell differentiation
MAPK signaling pathway
NF−kappa B signaling pathway
Hippo signaling pathway
Cytokine−cytokine receptor interaction
NOD−like receptor signaling pathway
Proteoglycans in cancer
Apoptosis
Synaptic vesicle cycle
Tight junction
Regulation of actin cytoskeleton
Spliceosome
Valine, leucine and isoleucine degradation
Ribosome biogenesis in eukaryotes
−log10(FDR)
C
Correlation
positive
negative
ns
−log10 (adj p.value)
R (protein vs RNA of each gene)
D
Positive
cell surface
collagen−containing extracellular matrix
extracellular matrix
external encapsulating structure
secretory granule
endoplasmic reticulum lumen
actin filament bundle
actin cytoskeleton
vesicle lumen
secretory granule lumen
actomyosin
vacuolar lumen
cytoplasmic vesicle lumen
stress fiber
contractile actin filament bundle
cell−cell junction
actin filament
lysosomal lumen
azurophil granule lumen
podosome
brush border
cell−substrate junction
blood microparticle
focal adhesion
primary lysosome
azurophil granule
cluster of actin−based cell projections
endoplasmic reticulum−Golgi intermediate compartment
adherens junction
ficolin−1−rich granule lumen
ficolin−1−rich granule
membrane raft
Gene Ratio
−log10(FDR)
NS
transferase complex
chromatin
mitochondrial protein−containing complex
nuclear body
ribosomal subunit
large ribosomal subunit
ubiquitin ligase complex
acetyltransferase complex
protein acetyltransferase complex
ribosome
histone acetyltransferase complex
transcription regulator complex
mitochondrial inner membrane
organelle inner membrane
RNA polymerase II transcription regulator complex
organellar ribosome
mitochondrial ribosome
E
R = 0.54, p < 2.2e−16
R = 0.47, p = 0.0074
R = 0.48, p < 2.2e−16
p = 0.0066
Protein abundance (signal/length)
RNA abundance (mean TPM)
Correlation
positive
negative
ns
F
Correlation
positive
ns
−log10 (adj p.value)
R (protein vs RNA of each gene)
G
R = 0.93, p < 2.2e−16
Mean Nanostring signals
Mean RNA−seq TPM
H
protein
RNA
Coefficient of Variation
Rank of RNA abundance (TPM)


Figure S4

**Fig. S4. Comparative analyses of human islet transcriptomes and proteomes. (A)** Correlation between the log2 fold changes of differentially expressed RNAs and proteins (T2D vs ND, adjusted $p < 0.05$). **(B)** Hallmark and selected KEGG gene sets enriched from the transcriptomic or proteomic changes in T2D (T2D vs ND, FDR<0.005 from transcriptome or proteome). **(C)** Within gene, across sample RNA-protein correlations among the 90 donor samples (82 ND and 8 T2D) and 7609 genes that were commonly measured in RNA-seq and proteomics. 1422 (19%) or 37 (0.5%) genes are positive or negatively correlated, respectively. **(D)** GO - Cellular Component (GO_CC) functional enrichment analysis of the gene sets enriched by the positive (top) or non-significant (bottom) RNA-protein correlations. (FDR<0.0001). **(E)** Across-gene correlation between RNAs and proteins among the genes that have positive, negative, or non-significant (ns) within-gene correlations in (C). Statistical difference between their correlation coefficient (R) were calculated using the cocor R package **(F)** Within gene, across sample correlation between RNAs measured by RNA-seq and NanoString. 82 out of the 99 commonly detected genes (83%) are positively correlated among the 64 common donor samples. **(G)** Across-gene correlation between RNAs measured by RNA-seq and NanoString. **(H)** Variance (Coefficient of Variation) of RNA and protein of each gene (ranked by the RNA abundance).

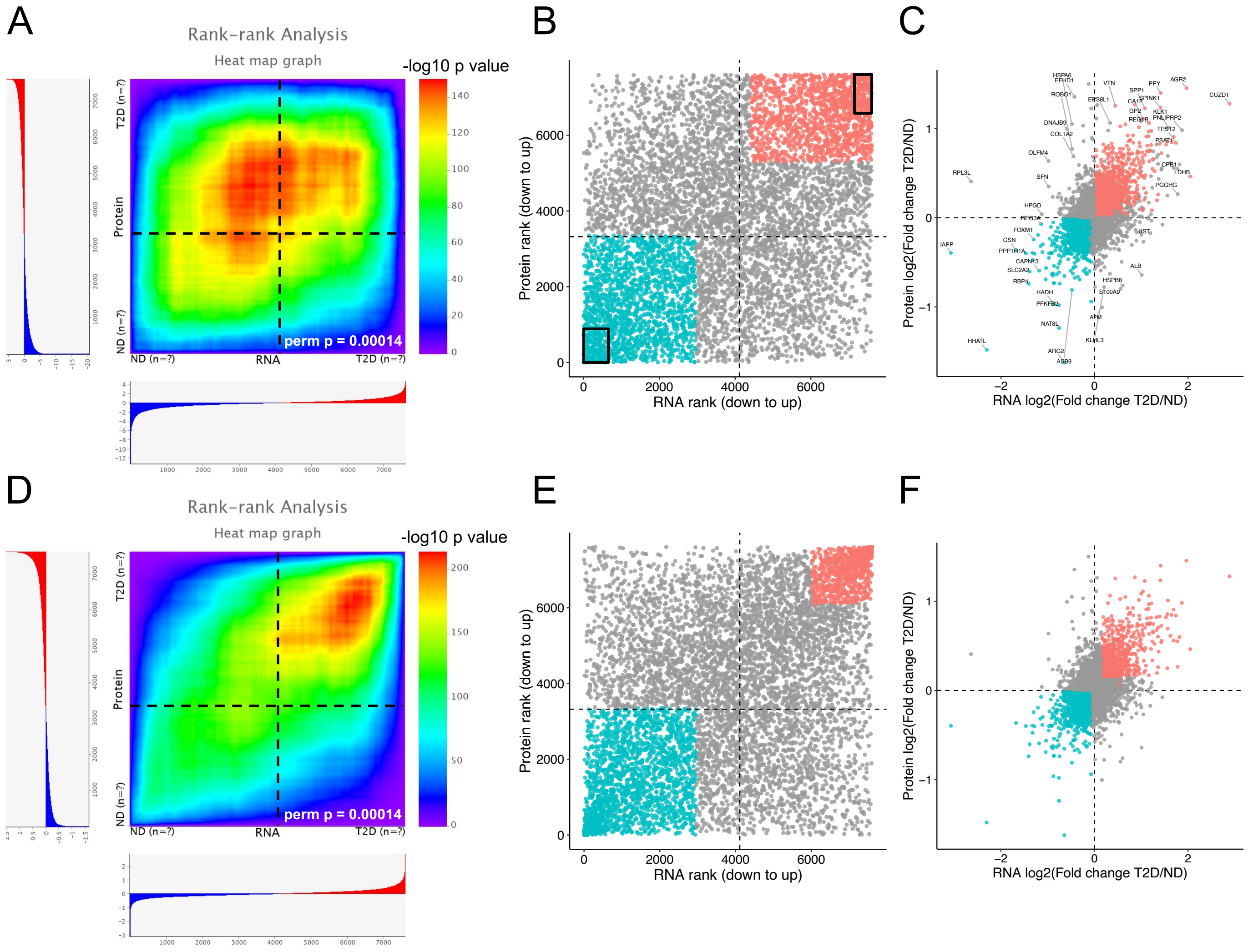


**Figure S5. Rank-rank hypergeometric overlap (RRHO) test comparing proteomic and transcriptomic changes in T2D.** Proteins and RNAs commonly detected in our proteomics and RNA-seq datasets were ranked by –log10 p value **(A-C)** or by log2 fold change **(D-F)** in our differential expression analysis (T2D/ND). **(A)** Hypergeometric overlap map showing the hypergeometric p values (adjusted by Benjamini-Yekutieli FDR) at each rank threshold. **(B)** Rank-Rank scatter plot with the most statistically significant overlapping gene set colored in pink (up-up) or blue (down-down). Proteins and RNAs within the black square were altered in T2D (using a less stringent cut off - unadjusted p value < 0.05 in differential expression analysis), which are only a small subset of the overlapping gene set determined by RRHO. **(C)** The log2 fold changes of proteins and RNAs with the most statistically significant overlapping gene set colored in pink (up-up) or blue (down-down). **(D-F)** Same as (A-C) but using log2 fold changes to rank the proteins and RNAs in RRHO test.

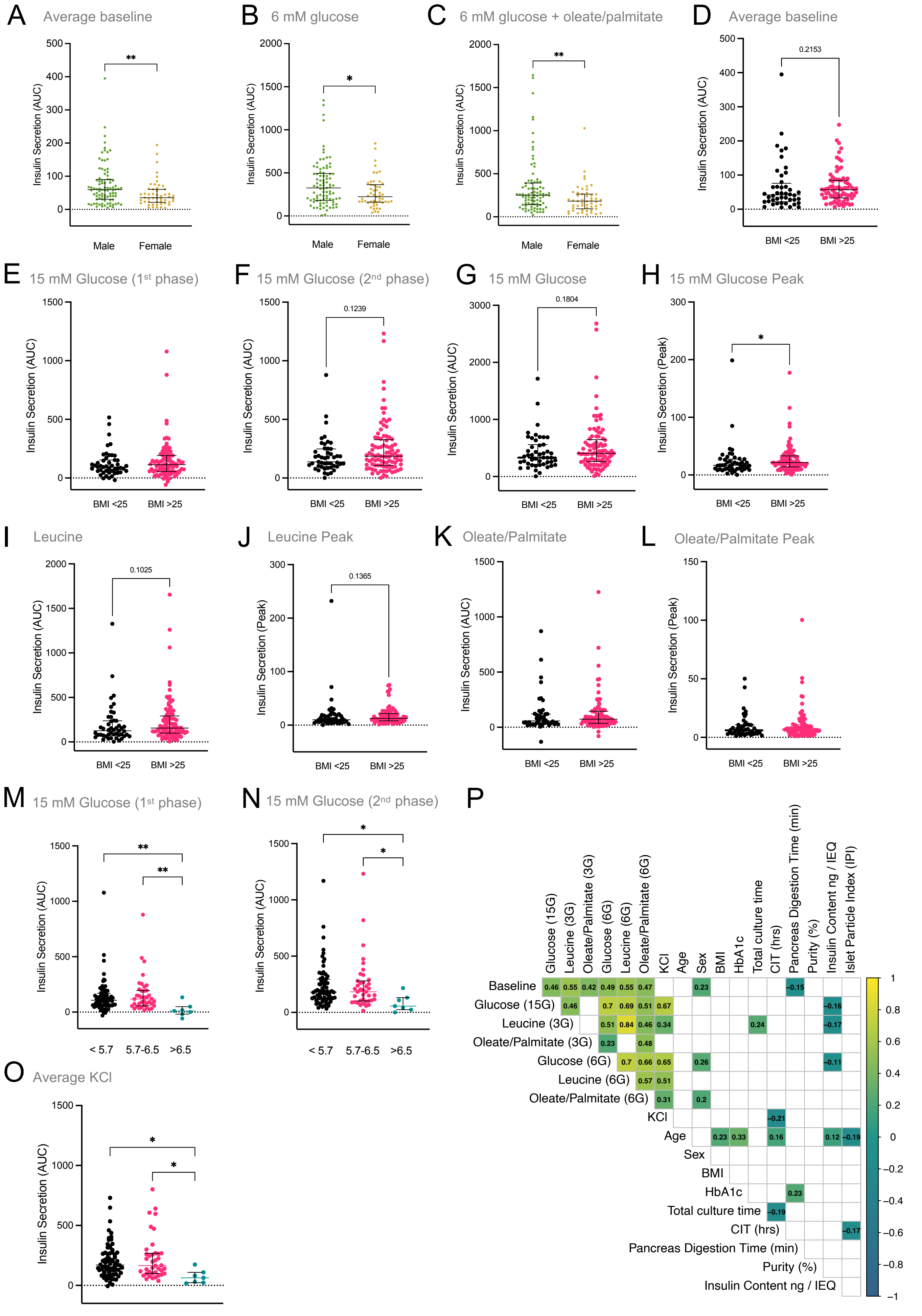
A Average baseline
Insulin Secretion (AUC)
Male
Female
**
B 6 mM glucose
*
C 6 mM glucose + oleate/palmitate
**
D Average baseline
0.2153
BMI <25
BMI >25
E 15 mM Glucose (1st phase)
F 15 mM Glucose (2nd phase)
0.1239
G 15 mM Glucose
0.1804
H 15 mM Glucose Peak
Insulin Secretion (Peak)
*
I Leucine
0.1025
J Leucine Peak
0.1365
K Oleate/Palmitate
L Oleate/Palmitate Peak
M 15 mM Glucose (1st phase)
< 5.7
5.7-6.5
>6.5
N 15 mM Glucose (2nd phase)
O Average KCl
P
Glucose (15G)
Leucine (3G)
Oleate/Palmitate (3G)
Glucose (6G)
Leucine (6G)
Oleate/Palmitate (6G)
KCl
Age
Sex
BMI
HbA1c
Total culture time
CIT (hrs)
Pancreas Digestion Time (min)
Purity (%)
Insulin Content ng / IEQ
Islet Particle Index (IPI)
Baseline 0.46 0.55 0.42 0.49 0.55 0.47 0.23 −0.15
Glucose (15G) 0.46 0.7 0.69 0.51 0.67 −0.16
Leucine (3G) 0.51 0.84 0.46 0.34 0.24 −0.17
Oleate/Palmitate (3G) 0.23 0.48
Glucose (6G) 0.7 0.66 0.65 0.26 −0.11
Leucine (6G) 0.57 0.51
Oleate/Palmitate (6G) 0.31 0.2
KCl −0.21
Age 0.23 0.33 0.16 0.12 −0.19
Sex
BMI
HbA1c 0.23
Total culture time −0.19
CIT (hrs) −0.17
Pancreas Digestion Time (min)
Purity (%)
Insulin Content ng / IEQ

Figure S6

**Fig. S6. Donor metrics and islet function. (A-C)** Shows direct comparisons of AUC values of indicated areas of the perifusion curve. between all donors separated by biological sex (male (n=89) and female (n=51)). Only measurements that show significant differences between the male and female donors are shown. **(D-L)** Shows direct comparisons of selected AUC values of indicated areas of the perifusion curve between all donors separated by BMI. **(M-O)** Shows direct comparisons of 1st and 2nd phase 15 mM glucose-stimulated insulin secretion as well as KCl-stimulated insulin secretion between all donors stratified by HbA1c. **(P)** Multiple comparisons matrix showing correlations between indicated parameters. Only significant R-values are shown.

A

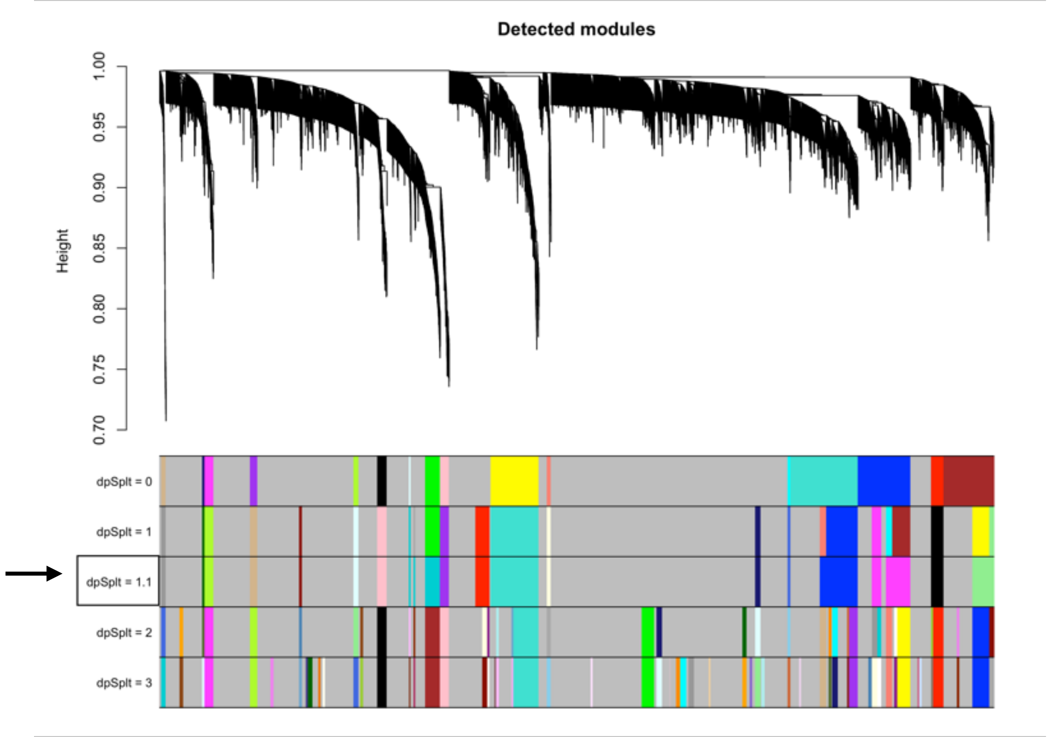
Detected modules
Height
1.00
0.95
0.90
0.85
0.80
0.75
0.70
dpSplt = 0
dpSplt = 1
dpSplt = 1.1
dpSplt = 2
dpSplt = 3

B

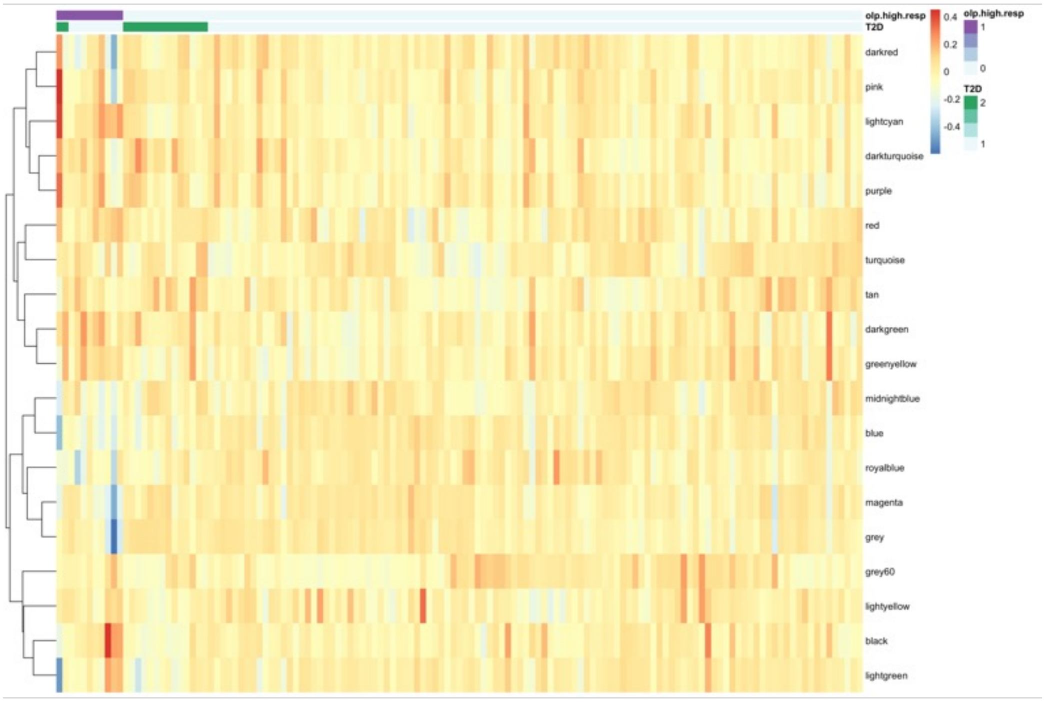
olp.high.resp
T2D
darkred
pink
lightcyan
darkturquoise
purple
red
turquoise
tan
darkgreen
greenyellow
midnightblue
blue
royalblue
magenta
grey
grey60
lightyellow
black
lightgreen
0.4
0.2
0
-0.2
-0.4
olp.high.resp
1
0
T2D
2
1

Figure S7

**Fig. S7. Detailed proteomics co-expression network results. (A)** Diagram showing hierarchically clustered proteins and their module membership. The height of the diagram corresponds to the dissimilarity of the two merged proteins for each node in the dendrogram. The bands below the dendrogram show detected modules for 4 different values of the 'deepSplit' parameter (dpSplit = 0, 1, 2, 3), as well as modules merged based on the correlation of their eigengenes for deepSplit parameter = 1 (dpSplit = 1.1; these module definitions were used for all downstream analyses). **(B)** Heatmap of module eigengenes for all donors. Donors are annotated with whether they were oleate palmitate high responders (purple = yes, grey = no) and whether they were diagnosed with type 2 diabetes (green = yes, grey = no). Modules (rows) are hierarchically clustered while donors (columns) are sorted first according to whether they are oleate palmitate high responders, and second according to whether they were reported to have type 2 diabetes .

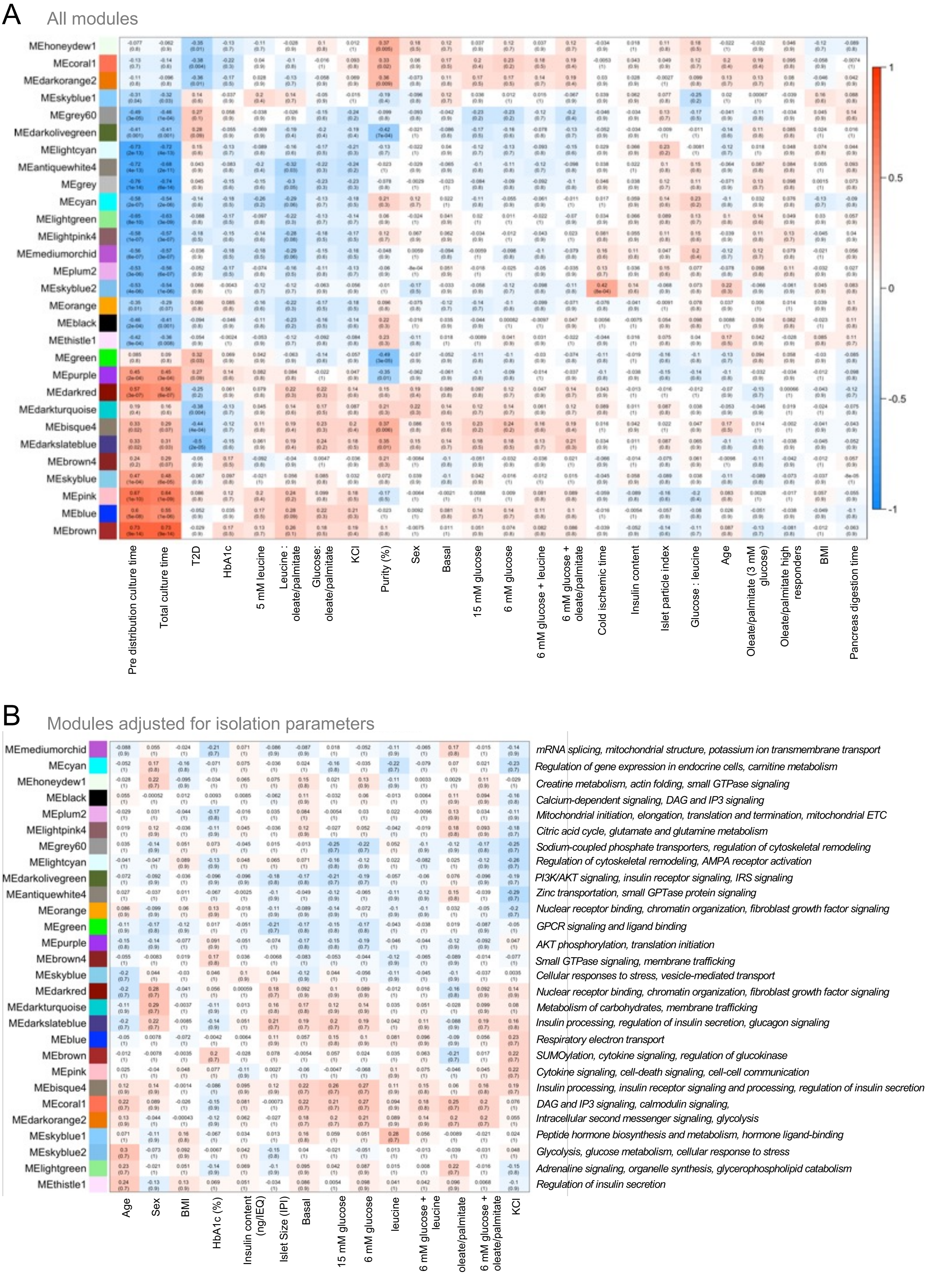
A
All modules
MEhoneydew1
MEcoral1
MEdarkorange2
MEskyblue1
MEgrey60
MEdarkolivegreen
MElightcyan
MEantiquewhite4
MEgrey
MEcyan
MElightgreen
MElightpink4
MEmediumorchid
MEplum2
MEskyblue2
MEorange
MEblack
MEthistle1
MEgreen
MEpurple
MEdarkred
MEdarkturquoise
MEbisque4
MEdarkslateblue
MEbrown4
MEskyblue
MEpink
MEblue
MEbrown
Pre distribution culture time
Total culture time
T2D
HbA1c
5 mM leucine
Leucine : oleate/palmitate
Glucose: oleate/palmitate
KCl
Purity (%)
Sex
Basal
15 mM glucose
6 mM glucose
6 mM glucose + leucine
6 mM glucose + oleate/palmitate
Cold ischemic time
Insulin content
Islet particle index
Glucose : leucine
Age
Oleate/palmitate (3 mM glucose)
Oleate/palmitate high responders
BMI
Pancreas digestion time
1
0.5
0
-0.5
-1
B
Modules adjusted for isolation parameters
MEmediumorchid
MEcyan
MEhoneydew1
MEblack
MEplum2
MElightpink4
MEgrey60
MElightcyan
MEdarkolivegreen
MEantiquewhite4
MEorange
MEgreen
MEpurple
MEbrown4
MEskyblue
MEdarkred
MEdarkturquoise
MEdarkslateblue
MEblue
MEbrown
MEpink
MEbisque4
MEcoral1
MEdarkorange2
MEskyblue1
MEskyblue2
MElightgreen
MEthistle1
Age
Sex
BMI
HbA1c (%)
Insulin content (ng/IEQ)
Islet Size (IPI)
Basal
15 mM glucose
6 mM glucose
leucine
6 mM glucose + leucine
oleate/palmitate
6 mM glucose + oleate/palmitate
KCl
mRNA splicing, mitochondrial structure, potassium ion transmembrane transport
Regulation of gene expression in endocrine cells, carnitine metabolism
Creatine metabolism, actin folding, small GTPase signaling
Calcium-dependent signaling, DAG and IP3 signaling
Mitochondrial initiation, elongation, translation and termination, mitochondrial ETC
Citric acid cycle, glutamate and glutamine metabolism
Sodium-coupled phosphate transporters, regulation of cytoskeletal remodeling
Regulation of cytoskeletal remodeling, AMPA receptor activation
PI3K/AKT signaling, insulin receptor signaling, IRS signaling
Zinc transportation, small GPTase protein signaling
Nuclear receptor binding, chromatin organization, fibroblast growth factor signaling
GPCR signaling and ligand binding
AKT phosphorylation, translation initiation
Small GTPase signaling, membrane trafficking
Cellular responses to stress, vesicle-mediated transport
Nuclear receptor binding, chromatin organization, fibroblast growth factor signaling
Metabolism of carbohydrates, membrane trafficking
Insulin processing, regulation of insulin secretion, glucagon signaling
Respiratory electron transport
SUMOylation, cytokine signaling, regulation of glucokinase
Cytokine signaling, cell-death signaling, cell-cell communication
Insulin processing, insulin receptor signaling and processing, regulation of insulin secretion
DAG and IP3 signaling, calmodulin signaling,
Intracellular second messenger signaling, glycolysis
Peptide hormone biosynthesis and metabolism, hormone ligand-binding
Glycolysis, glucose metabolism, cellular response to stress
Adrenaline signaling, organelle synthesis, glycerophospholipid catabolism
Regulation of insulin secretion

Figure S8

**Fig. S8. Association between RNAseq modules and donor metadata.** Heatmap of module - metadata associations, with pairwise correlations computed as **(A)** the Pearson correlation between module eigengenes and metadata values, and **(B)** the partial Pearson correlation between module eigengenes and metadata values other than technical isolation parameters (culture time, digestion time, cold ischemia time, and percent purity), with these parameters included as covariates in the partial correlation calculations. Cell color corresponds to correlation value, and each cell is annotated with the correlation value and FDR-adjusted p-value (FDR = 0.05) in brackets. Heatmap row (modules) and columns (metadata values) are hierarchically clustered based on correlation magnitude. Modules in B) are additionally labeled with representative functional themes from overrepresentation gene set analysis with the KEGG, Reactome, Gene Ontology Cellular Component, Molecular Function, and Biological Process functional libraries.

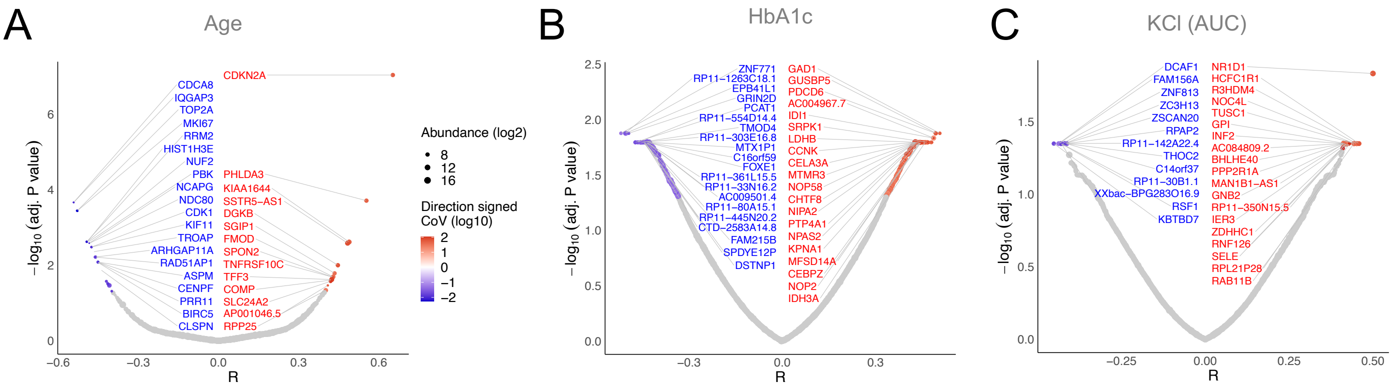


**Fig. S9. RNAseq versus donor metadata and islet function.** Volcano plots for mRNAs significantly negatively (blue) or positively (red) correlated to **(A)** donor age, **(B)** donor HbA1c, and **(C)** insulin secretion stimulated by direct depolarization with KCl. Protein abundances (log2) are depicted by size of circle, and the coefficient of variance (log10) is depicted by color gradient.

Figure S9

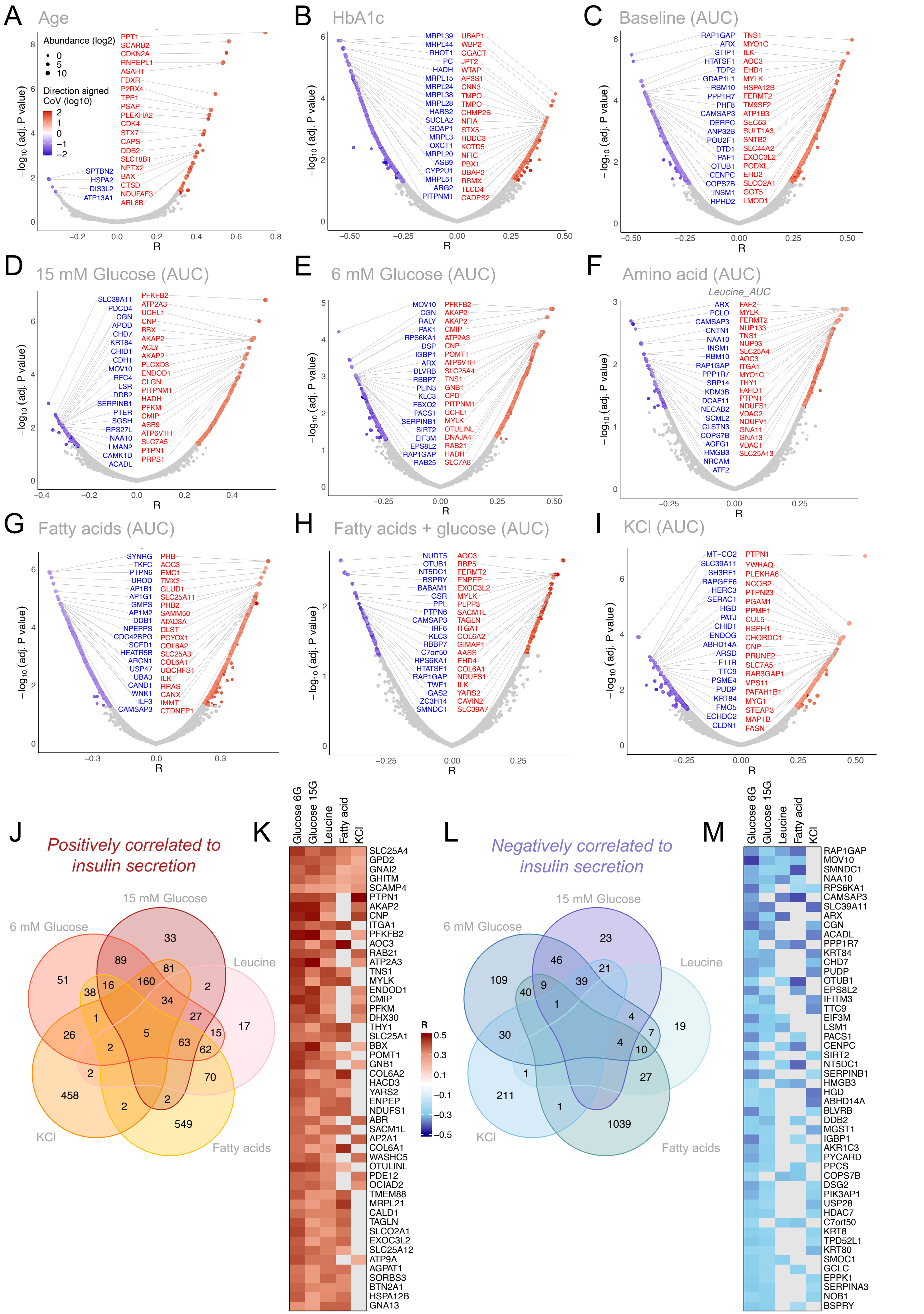

A Age
Abundance (log2)
0
5
10
Direction signed CoV (log10)
2
1
0
−1
−2
PPT1
SCARB2
CDKN2A
RNPEPL1
ASAH1
FDXR
P2RX4
TPP1
PSAP
PLEKHA2
CDK4
STX7
CAPS
DDB2
SLC18B1
NPTX2
BAX
CTSD
NDUFAF3
ARL8B
SPTBN2
HSPA2
DIS3L2
ATP13A1
−log10 (adj. P value)
R
B HbA1c
MRPL39
MRPL44
RHOT1
PC
HADH
MRPL15
MRPL24
MRPL38
MRPL28
HARS2
SUCLA2
GDAP1
MRPL3
OXCT1
MRPL20
ASB9
CYP2U1
MRPL51
ARG2
PITPNM1
UBAP1
WBP2
GGACT
JPT2
WTAP
AP3S1
CNN3
TMPO
TMPO
CHMP2B
NFIA
STX5
HDDC3
KCTD5
NFIC
PBX1
UBAP2
RBMX
TLCD4
CADPS2
C Baseline (AUC)
RAP1GAP
ARX
STIP1
HTATSF1
TDP2
GDAP1L1
RBM10
PPP1R7
PHF8
CAMSAP3
DERPC
ANP32B
POU2F1
DTD1
PAF1
OTUB1
CENPC
COPS7B
INSM1
RPRD2
TNS1
MYO1C
ILK
AOC3
EHD4
MYLK
HSPA12B
FERMT2
TM9SF2
ATP1B3
SEC63
SULT1A3
SNTB2
SLC44A2
EXOC3L2
PODXL
EHD2
SLCO2A1
GGT5
LMOD1
D 15 mM Glucose (AUC)
SLC39A11
PDCD4
CGN
APOD
CHD7
KRT84
CHID1
CDH1
MOV10
RFC4
LSR
DDB2
SERPINB1
PTER
SGSH
RPS27L
NAA10
LMAN2
CAMK1D
ACADL
PFKFB2
ATP2A3
UCHL1
CNP
BBX
AKAP2
ACLY
AKAP2
PLCXD3
ENDOD1
CLGN
PITPNM1
HADH
PFKM
CMIP
ASB9
ATP6V1H
SLC7A5
PTPN1
PRPS1
E 6 mM Glucose (AUC)
MOV10
CGN
RALY
PAK1
RPS6KA1
DSP
IGBP1
ARX
BLVRB
RBBP7
PLIN3
KLC3
FBXO2
PACS1
SERPINB1
SIRT2
EIF3M
EPS8L2
RAP1GAP
RAB25
PFKFB2
AKAP2
AKAP2
CMIP
ATP2A3
CNP
POMT1
ATP6V1H
SLC25A4
TNS1
GNB1
CPD
PITPNM1
UCHL1
MYLK
OTULINL
DNAJA4
RAB21
HADH
SLC7A8
F Amino acid (AUC)
Leucine_AUC
ARX
PCLO
CAMSAP3
CNTN1
NAA10
INSM1
RBM10
RAP1GAP
PPP1R7
SRP14
KDM3B
DCAF11
NECAB2
SCML2
CLSTN3
COPS7B
AGFG1
HMGB3
NRCAM
ATF2
FAF2
MYLK
FERMT2
NUP133
TNS1
NUP93
SLC25A4
AOC3
ITGA1
MYO1C
THY1
FAHD1
PTPN1
NDUFS1
VDAC2
NDUFV1
GNA11
GNA13
VDAC1
SLC25A13
G Fatty acids (AUC)
SYNRG
TKFC
PTPN6
UROD
AP1B1
AP1G1
GMPS
AP1M2
DDB1
NPEPPS
CDC42BPG
SCFD1
HEATR5B
ARCN1
USP47
UBA3
CAND1
WNK1
ILF3
CAMSAP3
PHB
AOC3
EMC1
TMX3
GLUD1
SLC25A11
PHB2
SAMM50
ATAD3A
DLST
PCYOX1
COL6A2
SLC25A3
COL6A1
UQCRFS1
ILK
RRAS
CANX
IMMT
CTDNEP1
H Fatty acids + glucose (AUC)
NUDT5
OTUB1
NT5DC1
BSPRY
BABAM1
GSR
PPL
PTPN6
CAMSAP3
IRF6
KLC3
RBBP7
C7orf50
RPS6KA1
HTATSF1
RAP1GAP
TWF1
GAS2
ZC3H14
SMNDC1
AOC3
RBP5
FERMT2
ENPEP
EXOC3L2
MYLK
PLPP3
SACM1L
TAGLN
ITGA1
COL6A2
GIMAP1
AASS
EHD4
COL6A1
NDUFS1
ILK
YARS2
CAVIN2
SLC39A7
I KCl (AUC)
MT−CO2
SLC39A11
SH3RF1
RAPGEF6
HERC3
SERAC1
HGD
PATJ
CHID1
ENDOG
ABHD14A
ARSD
F11R
TTC9
PSME4
PUDP
KRT84
FMO5
ECHDC2
CLDN1
PTPN1
YWHAQ
PLEKHA6
NCOR2
PTPN23
PGAM1
PPME1
CUL5
HSPH1
CHORDC1
CNP
PRUNE2
SLC7A5
RAB3GAP1
VPS11
PAFAH1B1
MYG1
STEAP3
MAP1B
FASN
J Positively correlated to insulin secretion
15 mM Glucose
6 mM Glucose
Leucine
KCl
Fatty acids
K
Glucose 6G
Glucose 15G
Leucine
Fatty acid
KCl
SLC25A4
GPD2
GNAI2
GHITM
SCAMP4
PTPN1
AKAP2
CNP
ITGA1
PFKFB2
AOC3
RAB21
ATP2A3
TNS1
MYLK
ENDOD1
CMIP
PFKM
DHX30
THY1
SLC25A1
BBX
POMT1
GNB1
COL6A2
HACD3
YARS2
ENPEP
NDUFS1
ABR
SACM1L
AP2A1
COL6A1
WASHC5
OTULINL
PDE12
OCIAD2
TMEM88
MRPL21
CALD1
TAGLN
SLCO2A1
EXOC3L2
SLC25A12
ATP9A
AGPAT1
SORBS3
BTN2A1
HSPA12B
GNA13
R
0.5
0.3
0.1
−0.1
−0.3
−0.5
L Negatively correlated to insulin secretion
15 mM Glucose
6 mM Glucose
Leucine
KCl
Fatty acids
M
Glucose 6G
Glucose 15G
Leucine
Fatty acid
KCl
RAP1GAP
MOV10
SMNDC1
NAA10
RPS6KA1
CAMSAP3
SLC39A11
ARX
CGN
ACADL
PPP1R7
KRT84
CHD7
PUDP
OTUB1
EPS8L2
IFITM3
TTC9
EIF3M
LSM1
PACS1
CENPC
SIRT2
NT5DC1
SERPINB1
HMGB3
HGD
ABHD14A
BLVRB
DDB2
MGST1
IGBP1
AKR1C3
PYCARD
PPCS
COPS7B
DSG2
PIK3AP1
USP28
HDAC7
C7orf50
KRT8
TPD52L1
KRT80
SMOC1
GCLC
EPPK1
SERPINA3
NOB1
BSPRY


Figure S10

**Fig. S10. Correlationa of individual proteins and mRNAs with insulin secretory responses.** Volcano plots are shown depicting significant positive (red) and negative (blue) correlations of proteins to continuous donor characteristics: **(A)** donor age and **(B)** HbA1c; or functional parameters in response to: **(C)** 3 mM glucose **(D)** 15 mM glucose **(E)** 6 mM glucose **(F)** 5 mM leucine **(G)** 1.5 mM oleate/palmitate (1:1 molar ratio) **(H)** 1.5 mM oleate/palmitate + 6 mM glucose and **(I)** 30 mM KCl. Protein abundances (log2) are depicted by size of circle, and the coefficient of variance (log10) is depicted by color gradient. **(J)** Venn diagram showing the overlap of the number of protein abundances that positively correlate to the indicated nutrient stimuli. **(K)** Heat map depicts the top 50 most positively correlative proteins to the indicated stimuli. **(L-M)** Shows the same as (J-K), but for negative correlations.

Age

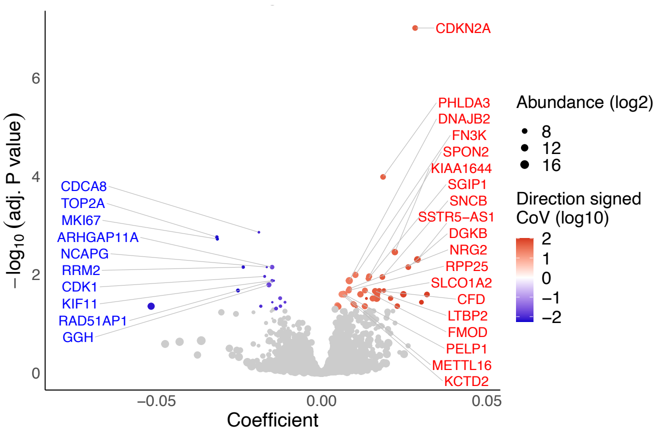


**Fig. S11. Association between RNAseq and donor metadata.** Volcano plot depicting significant positive (red) and negative (blue) linear regression coefficients (adjusting for T2D status) of transcripts associated with donor age. RNA expression (abundances) are depicted by size of circle (log2), and the coefficient of variation (log10) is depicted by color gradient.

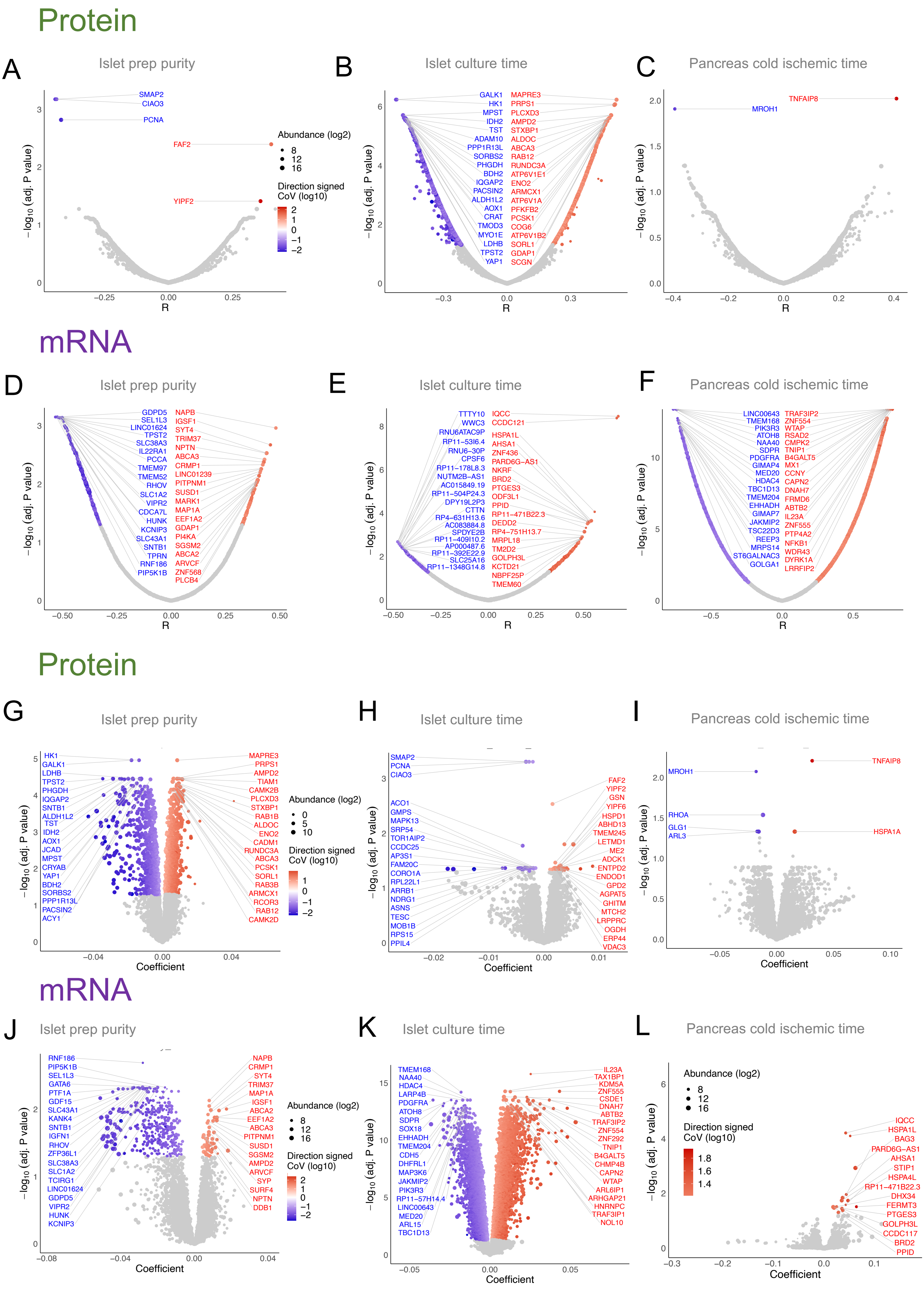
Protein
A Islet prep purity
B Islet culture time
C Pancreas cold ischemic time
mRNA
D Islet prep purity
E Islet culture time
F Pancreas cold ischemic time
Protein
G Islet prep purity
H Islet culture time
I Pancreas cold ischemic time
mRNA
J Islet prep purity
K Islet culture time
L Pancreas cold ischemic time
Abundance (log2)
Direction signed CoV (log10)
−log10 (adj. P value)
R
Coefficient

Figure S12

**Fig. S12. Proteome and transcriptome versus islet isolation parameters.** Volcano plots for proteins significantly negatively (blue) or positively (red) correlated to **(A)** islet preparation purity, **(B)** total islet culture time and **(C)** cold ischemic time of the pancreas. **(D-F)** Shows the same as (A-C) except for correlations with the transcriptome. Protein abundances (log2) are depicted by size of circle, and the coefficient of variance (log10) is depicted by color gradient. All donors (independent of diabetes diagnosis) were used in these analyses. **(G-H)** depicts volcano plots with significant positive (red) and negative (blue) linear regression coefficients (adjusting for T2D status) of proteins to islet preparation purity, total islet culture time and cold ischemic time of the pancreas. **(J-L)** Shows the same as (G-I) except for associations with the transcriptome. Protein and transcript abundances (log2) are depicted by size of circle, and the coefficient of variation (log10) is depicted by color gradient.

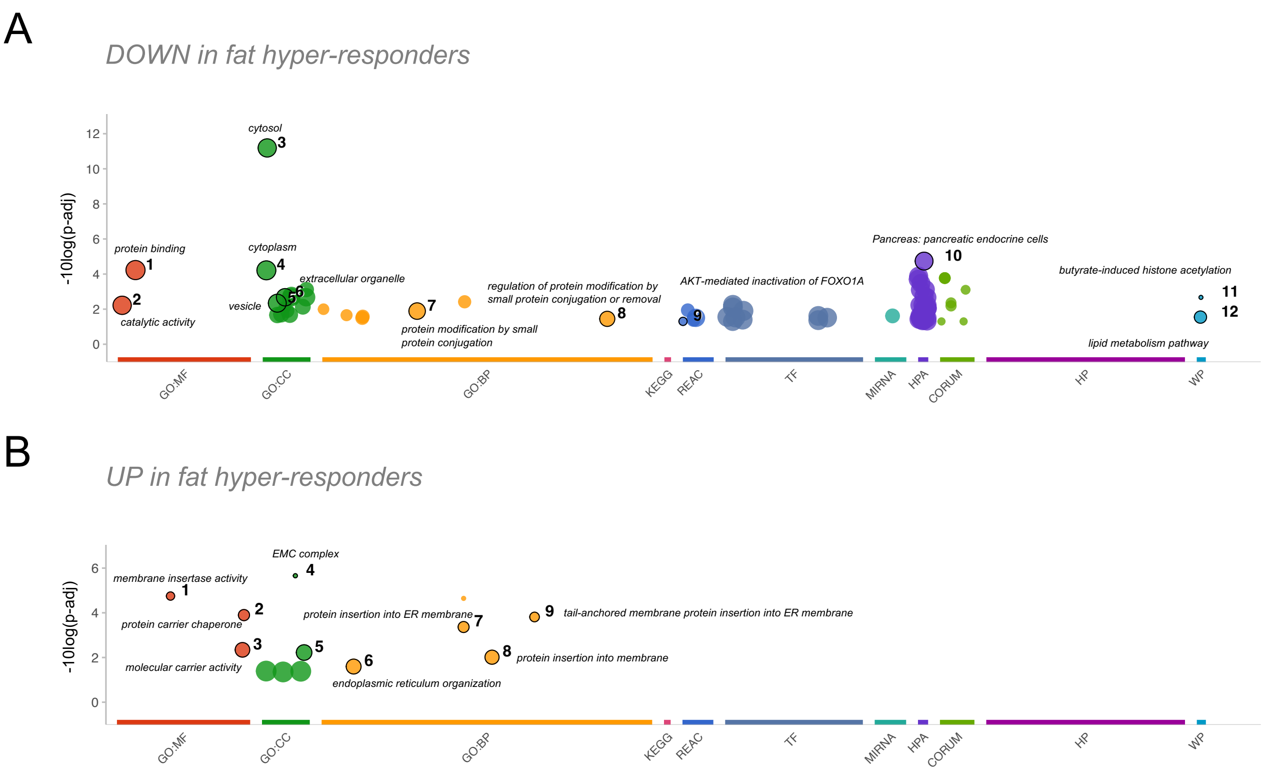


**Fig. S13. Pathway analysis of differentially abundant proteins in fat hyper-responders. (A)** Pathway analysis of proteins that are less abundant in fat hyper-responsive donors showing enriched biological functions and pathways obtained from multiple databases including (among others), Gene Ontology (GO), KEGG and Reactome. **(B)** Shows the same as (A) except for proteins that are more abundant in fat hyper-responsive donors.

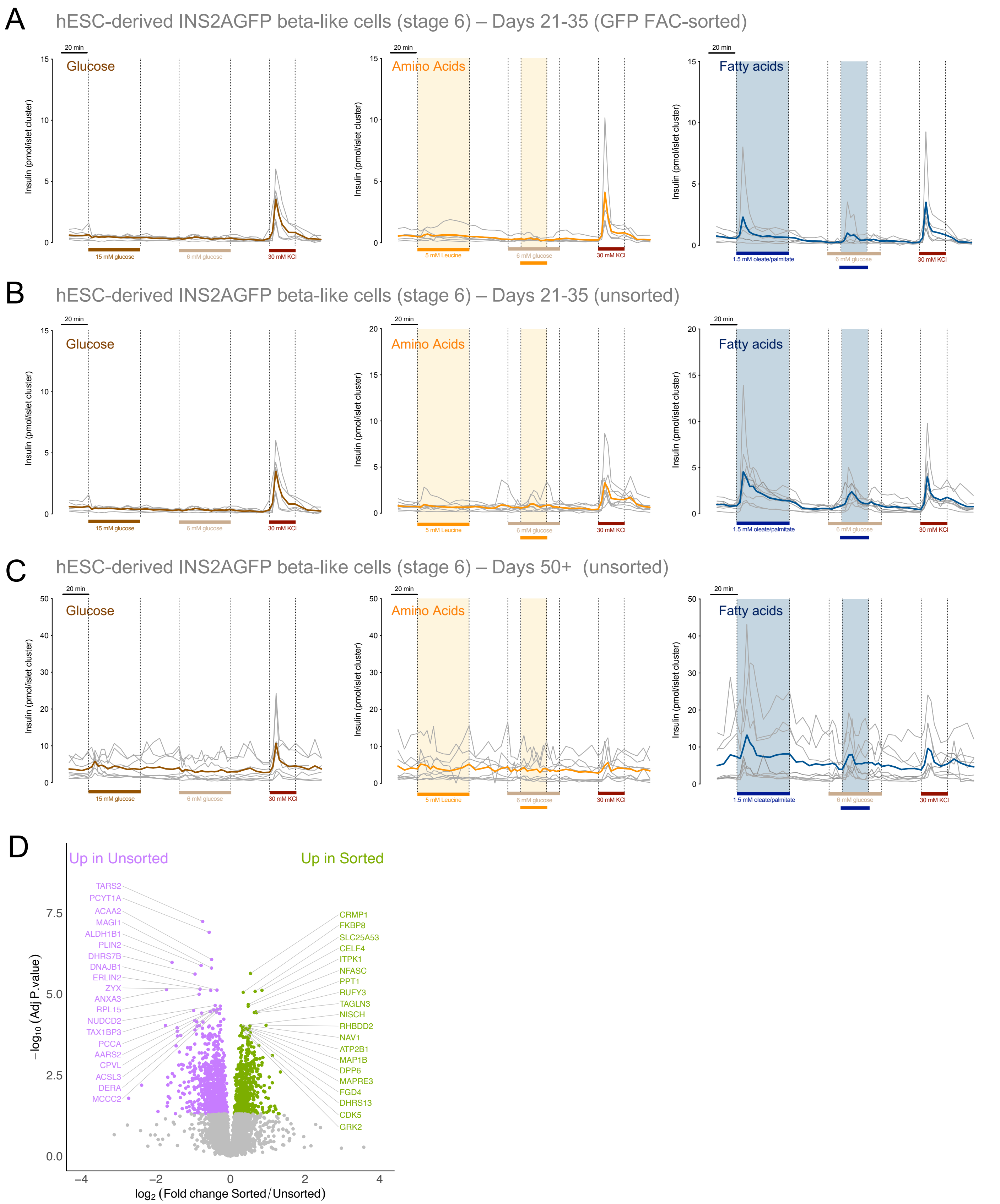
A
hESC-derived INS2AGFP beta-like cells (stage 6) – Days 21-35 (GFP FAC-sorted)
B
hESC-derived INS2AGFP beta-like cells (stage 6) – Days 21-35 (unsorted)
C
hESC-derived INS2AGFP beta-like cells (stage 6) – Days 50+ (unsorted)
20 min
Glucose
Amino Acids
Fatty acids
Insulin (pmol/islet cluster)
15 mM glucose
6 mM glucose
30 mM KCl
5 mM Leucine
1.5 mM oleate/palmitate
D
Up in Unsorted
Up in Sorted
TARS2
PCYT1A
ACAA2
MAGI1
ALDH1B1
PLIN2
DHRS7B
DNAJB1
ERLIN2
ZYX
ANXA3
RPL15
NUDCD2
TAX1BP3
PCCA
AARS2
CPVL
ACSL3
DERA
MCCC2
CRMP1
FKBP8
SLC25A53
CELF4
ITPK1
NFASC
PPT1
RUFY3
TAGLN3
NISCH
RHBDD2
NAV1
ATP2B1
MAP1B
DPP6
MAPRE3
FGD4
DHRS13
CDK5
GRK2
−log10 (Adj P.value)
log2 (Fold change Sorted / Unsorted)

Figure S14

**Fig. S14. Stem cell derived islet cluster individual response traces. (A)** Individual (grey) and averaged traces (colored) of dynamic insulin secretion measurements in response to glucose (15 or 6 mM), leucine (5 mM), 1.5 mM oleate and palmitate (1:1 mixture) or 30 mM KCl in stem cell derived islet clusters (5 differentiations, GFP-Fluorescence-activated cell (FAC) sorted, stage 6 (days 21-25). Basal glucose was 3 mM. **(B)** Same as (A) except in 8 differentiations of unsorted clusters (stage 6, days 21-30). **(C)** Same as (B) except in 8 differentiations of older-maturing clusters (stage 6, days 55+). **(D)** Volcano plot showing differential protein abundance between GFP-FAC sorted stem cell derived islet like clusters (purple, n=11) and GFP-unsorted clusters (green, n=25). The top 30 most significant differentially abundant proteins are highlighted by labelling with gene name.

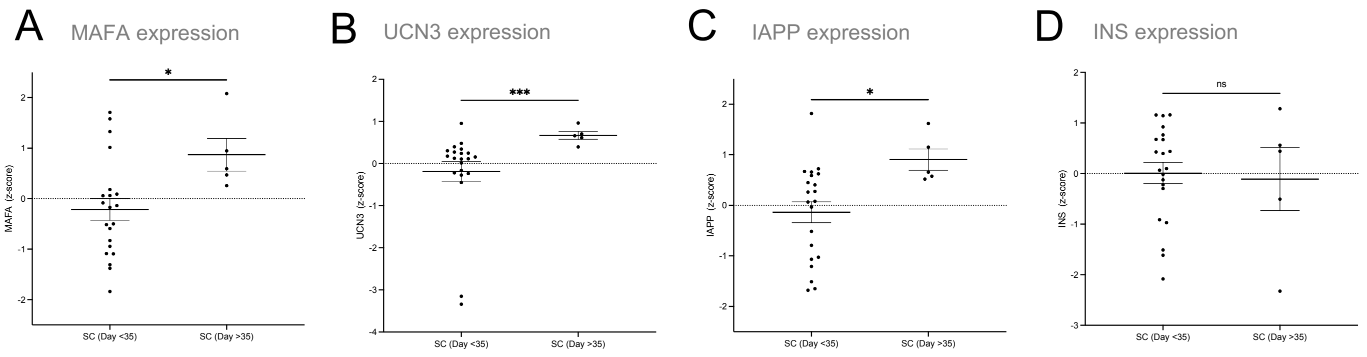


**Fig. S15. Transcriptional maturation of stem cell derived islet-like clusters.** Calculated z-scores of key maturation transcripts **(A)** MAFA, **(B)** UCN3, **(C)** IAPP and **(D)** INS are compared between stem cell-derived islet clusters less than and greater than 35 days of age.
